# Fine-mapping genomic loci refines bipolar disorder risk genes

**DOI:** 10.1101/2024.02.12.24302716

**Authors:** Maria Koromina, Ashvin Ravi, Georgia Panagiotaropoulou, Brian M. Schilder, Jack Humphrey, Alice Braun, Tim Bidgeli, Chris Chatzinakos, Brandon Coombes, Jaeyoung Kim, Xiaoxi Liu, Chikashi Terao, Kevin S. O.’Connell, Mark Adams, Rolf Adolfsson, Martin Alda, Lars Alfredsson, Till F. M. Andlauer, Ole A. Andreassen, Anastasia Antoniou, Bernhard T. Baune, Susanne Bengesser, Joanna Biernacka, Michael Boehnke, Rosa Bosch, Murray J. Cairns, Vaughan J. Carr, Miquel Casas, Stanley Catts, Sven Cichon, Aiden Corvin, Nicholas Craddock, Konstantinos Dafnas, Nina Dalkner, Udo Dannlowski, Franziska Degenhardt, Arianna Di Florio, Dimitris Dikeos, Frederike Tabea Fellendorf, Panagiotis Ferentinos, Andreas J. Forstner, Liz Forty, Mark Frye, Janice M. Fullerton, Micha Gawlik, Ian R. Gizer, Katherine Gordon-Smith, Melissa J. Green, Maria Grigoroiu-Serbanescu, José Guzman-Parra, Tim Hahn, Frans Henskens, Jan Hillert, Assen V. Jablensky, Lisa Jones, Ian Jones, Lina Jonsson, John R. Kelsoe, Tilo Kircher, George Kirov, Sarah Kittel-Schneider, Manolis Kogevinas, Mikael Landén, Marion Leboyer, Melanie Lenger, Jolanta Lissowska, Christine Lochner, Carmel Loughland, Donald MacIntyre, Nicholas G. Martin, Eirini Maratou, Carol A. Mathews, Fermin Mayoral, Susan L. McElroy, Nathaniel W. McGregor, Andrew McIntosh, Andrew McQuillin, Patricia Michie, Philip B. Mitchell, Paraskevi Moutsatsou, Bryan Mowry, Bertram Müller-Myhsok, Richard M. Myers, Igor Nenadić, Caroline Nievergelt, Markus M. Nöthen, John Nurnberger, Michael O.’Donovan, Claire O’Donovan, Roel A. Ophoff, Michael J. Owen, Christos Pantelis, Carlos Pato, Michele T. Pato, George P. Patrinos, Joanna M. Pawlak, Roy H. Perlis, Evgenia Porichi, Danielle Posthuma, Josep Antoni Ramos-Quiroga, Andreas Reif, Eva Z. Reininghaus, Marta Ribasés, Marcella Rietschel, Ulrich Schall, Peter R. Schofield, Thomas G. Schulze, Laura Scott, Rodney J. Scott, Alessandro Serretti, Cynthia Shannon Weickert, Jordan W. Smoller, Maria Soler Artigas, Dan J. Stein, Fabian Streit, Claudio Toma, Paul Tooney, Marquis P. Vawter, Eduard Vieta, John B. Vincent, Irwin D. Waldman, Thomas Weickert, Stephanie H. Witt, Kyung Sue Hong, Masashi Ikeda, Nakao Iwata, Beata Świątkowska, Hong-Hee Won, Howard J. Edenberg, Stephan Ripke, Towfique Raj, Jonathan R. I. Coleman, Niamh Mullins

## Abstract

Bipolar disorder (BD) is a heritable mental illness with complex etiology. While the largest published genome-wide association study identified 64 BD risk loci, the causal SNPs and genes within these loci remain unknown. We applied a suite of statistical and functional fine-mapping methods to these loci, and prioritized 17 likely causal SNPs for BD. We mapped these SNPs to genes, and investigated their likely functional consequences by integrating variant annotations, brain cell-type epigenomic annotations, brain quantitative trait loci, and results from rare variant exome sequencing in BD. Convergent lines of evidence supported the roles of genes involved in neurotransmission and neurodevelopment including *SCN2A, TRANK1, DCLK3, INSYN2B, SYNE1, THSD7A, CACNA1B, TUBBP5, PLCB3, PRDX5, KCNK4, CRTC3, AP001453*.*3, TRPT1, FKBP2, DNAJC4, RASGRP1, FURIN, FES, DPH1, GSDMB, MED24* and *THRA* in BD. These represent promising candidates for functional experiments to understand biological mechanisms and therapeutic potential. Additionally, we demonstrated that fine-mapping effect sizes can improve performance of BD polygenic risk scores across diverse populations, and present a high-throughput fine-mapping pipeline (https://github.com/mkoromina/SAFFARI).

## Introduction

Bipolar disorder (BD) is a heritable mental illness with complex etiology^1^. Heritability estimates from twin studies range between 60% and 90%^2–4^, while SNP-based heritability (h^2^_SNP_) calculations suggest that common genetic variants can explain up to 20% of the phenotypic variance of BD^5^. Genome-wide association studies (GWAS) of common variants have been successful in identifying associated genetic risk loci for BD^5–15^. For example, the largest published BD GWAS to date, conducted by the Psychiatric Genomics Consortium (PGC), comprised more than 40,000 BD cases and 370,000 controls from 57 cohorts of European ancestries, and identified 64 genome-wide significant (GWS) risk loci^16^. However, identifying the causal SNPs within these loci (i.e., SNPs responsible for the association signal at a locus and with a biological effect on the phenotype, as opposed to those associated due to linkage disequilibrium (LD) with a causal variant) is a major challenge.

Computational fine-mapping methods aim to identify independent causal variants within a genomic locus by modeling LD structure, SNP association statistics, number of causal variants, and/or prior probabilities of causality based on functional annotations. There are a variety of fine-mapping models ranging from regression to Bayesian methods, with different strengths and limitations^17–19^. For example, the Sum of Single Effects (SuSiE) model uses iterative Bayesian selection with posterior probabilities^20^, FINEMAP employs a stochastic search algorithm for SNP combinations^21^, and POLYgenic FUNctionally-informed fine-mapping (PolyFun) computes functional priors to improve fine-mapping accuracy^18^. Bayesian fine-mapping methods typically generate a posterior inclusion probability (PIP) of causality per SNP, and “credible sets” of SNPs, which represent the minimum set of SNPs with a specified probability of including the causal variant(s). Many methods can assume one or multiple causal variants per locus, and can now be applied to GWAS summary statistics from large and well-powered studies. This is highly advantageous for fine-mapping GWAS meta-analyses; however, the specification of appropriate LD structure is crucial for accurate fine-mapping. When LD cannot be obtained from the original cohort(s) (e.g. due to data access restrictions), it should instead be obtained from a sufficiently large sample that is ancestrally similar to the GWAS population^22^.

Fine-mapping methods have recently been applied to GWAS of psychiatric disorders. For example, a recent study using FINEMAP and integrating functional genomic data identified more than 100 genes likely to underpin associations in risk loci for schizophrenia^23^. Several fine-mapped candidates had particularly strong support for their pathogenic role in schizophrenia, due to convergence with rare variant associations^23^. Here, we use a suite of tools to conduct statistical and functional fine-mapping of 64 GWS risk loci for BD^16^ and assess the impact of the LD reference panel and fine-mapping window specifications. We link the likely causal SNPs to their relevant genes and investigate their potential functional consequences, by integrating functional genomic data, including brain cell-type-specific epigenomic annotations, and quantitative trait loci data. We also fine-mapped the major histocompatibility complex (MHC) separately by imputing human leukocyte antigen (HLA) variants, and assessed the impact of fine-mapping on polygenic risk score (PRS) predictions. Finally, we present a comprehensive fine-mapping pipeline implemented via Snakemake^24^ as a rapid, scalable, and cost-effective approach to prioritize likely variants from GWS risk loci. This strategy yielded promising candidate genes for future experiments to understand the mechanisms by which they increase risk of BD.

## Methods

### GWAS summary statistics and BD risk loci

Summary statistics from the latest published BD GWAS by the Psychiatric Genomics Consortium (PGC) were used as input to the fine-mapping pipeline^16^. Briefly, this GWAS comprised 41,917 BD cases, and 371,549 controls of European (EUR) ancestries, from 57 cohorts (**Supplemental Table S1**). Of these cohorts, 53 were imputed using the Haplotype Reference Consortium (HRC) EUR ancestry reference panel v1.0^25^. GWAS summary statistics were cleaned using DENTIST software^26^ yielding a total of 7,598,903 SNPs. The GWAS meta-analysis identified 64 independent loci associated with BD at GWS, which were selected for fine-mapping. Each GWS locus window was established around the GWS significant “top lead” SNP (*P* < 5 × 10^−8^), with boundaries defined by the positions of the 3’-most and 5’-most SNPs, requiring an LD r^2^ > 0.1 with the top lead SNP within a 3 Mb range, according to the LD structure of the HRC EUR reference panel^16^. Due to the complexity and long-range LD of the MHC/HLA region, this locus was analyzed separately (see section ‘Fine-mapping the MHC locus’). **Supplemental Table S2** shows the top lead SNP from each GWS locus, association statistics, locus boundaries, locus size, and locus names (as defined in the original GWAS)^16^. Excluding the MHC, GWS locus windows ranged between 14,960 - 3,730,000 bp in size.

### Conditional analysis

Figure 1. shows an overview of the fine-mapping pipeline. First, stepwise conditional analyses were conducted using GCTA-COJO^27^ to identify potential independent association signals within each locus, according to the LD structure of the HRC EUR reference panel. Association tests were performed for all SNPs in each locus, conditioning sequentially on the top lead SNP, until no conditional tests were significant (conditional P > 5 × 10^−6^), to calculate the number of independent association signals per locus. A less stringent P value threshold (P < 5 × 10^−6^) for significance was selected for the conditional analysis, consistent with the recommendations of Yang et al^27^.

**Figure 1.**
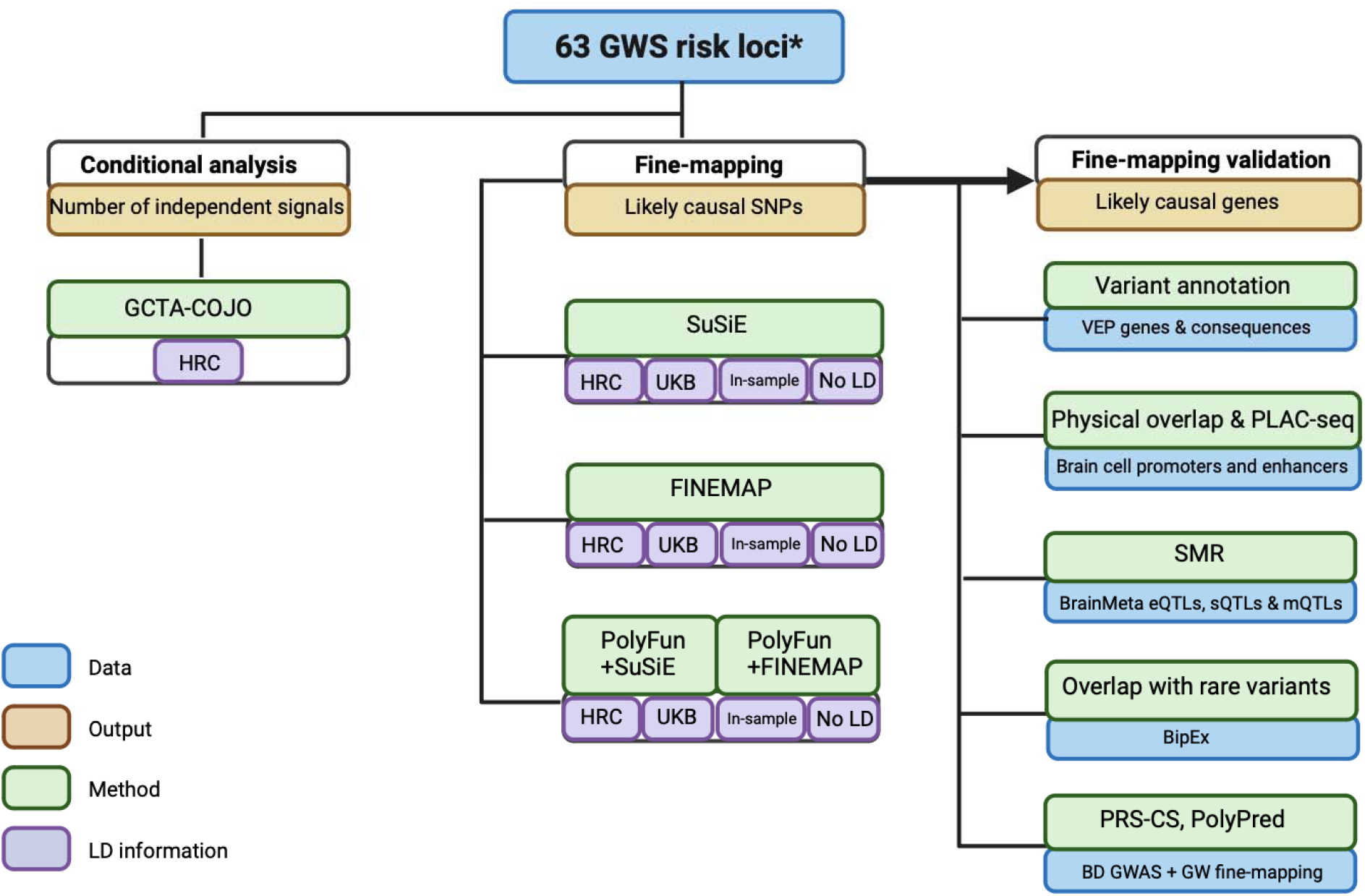
Schematic workflow of the fine-mapping pipeline developed for BD GWAS risk loci. Conditional analyses were performed within GWS loci using GCTA-COJO, based on the linkage disequilibrium (LD) structure of the Haplotype Reference Consortium (HRC) reference panel. Fine-mapping was conducted using statistical (SuSiE and FINEMAP) and functionally-informed (PolyFun) methods, according to the LD structure of the HRC, UK Biobank (UKB), and a subset of the GWAS data (“in-sample LD”), as well as implementing single-variant (no LD) fine-mapping. PolyFun functional priors were based on the published baseline-LF2.2 UKB model^28^. Fine-mapping results were validated computationally via Variant Effect Predictor (VEP) annotations and functional consequences, overlap with epigenomic peaks from brain cell-types, Summary-data-based Mendelian Randomization analysis (SMR) with brain expression, splicing and methylation QTL data, convergence with rare variant associations from the Bipolar Exome Sequencing Collaboration (BipEx), and testing whether fine-mapping effect sizes improve polygenic risk scores (PRS-CS and PolyPred). *The major histocompatibility complex (MHC) was fine-mapped using separate procedures (see section ‘Fine-mapping the MHC locus’).

### LD reference panels

Statistical and functional fine-mapping methods require information on LD between variants and selection of a genomic region (“window”) to fine-map. To examine the impact of LD on fine-mapping, analyses were performed using LD information from the HRC EUR reference panel, published LD matrices based on EUR ancestry individuals in the UK Biobank (UKB)^18^, and “in-sample” LD calculated from a subset of 48 BD cohorts in the PGC BD GWAS for which individual-level genetic data were available within the PGC (33,781 cases, 53,869 controls, all of EUR ancestries), representing 73% of the total effective sample size of the GWAS. Briefly, HRC-imputed dosage data were converted to hard calls with a genotype call probability cut-off of 0.8 and PLINK binary files were merged across cohorts, restricting to the set of unrelated individuals included in the GWAS, using PLINK v1.90^29^. Missingness rates per SNP were calculated in each cohort, and SNPs absent in all individuals from any one cohort were excluded from the merged dataset, yielding 7,594,494 SNPs overlapping with the GWAS summary statistics. Individual-level genetic data per chromosome were used as an “in-sample” LD reference panel for fine-mapping. We also performed single variant fine-mapping without any LD.

### Statistical and functional fine-mapping

GWS loci were fine-mapped using a suite of Bayesian fine-mapping methods that can be applied to GWAS summary statistics: SuSiE, FINEMAP, PolyFun+SuSiE, PolyFun+FINEMAP (**Figure 1**). SuSiE and FINEMAP are statistical fine-mapping methods, while PolyFun incorporates functional annotations as prior probabilities to improve subsequent fine-mapping accuracy^18,20,21^. Since these methods have different underlying assumptions, strengths and limitations, results were compared to examine convergence of evidence across methods. Briefly, each Bayesian method generates SNP-wise posterior inclusion probabilities of causality (PIP), and a 95% credible set (95% CS), defined as the minimum subset of SNPs that cumulatively have at least 95% probability of containing the causal SNP(s). PIP refers to the marginal probability that a SNP is included in any causal model, conditional on the observed data, hence providing weight of evidence that a SNP should be considered potentially causal.

First, single variant fine-mapping, which makes the simple assumption of one causal variant per locus (K = 1) and does not require LD information^18,20,21^, was performed within each GWS locus fine-mapping window. FINEMAP and SuSiE can assume multiple causal variants per locus, modeling the LD structure between them. Fine-mapping was additionally performed assuming the default maximum of five causal variants per locus (K = 5) and separately using the HRC, UKB and “in-sample” LD structures. Finally, PolyFun was used to incorporate 187 published functional annotations from the baseline-LF 2.2.UKB model^28^ to compute prior causal probabilities (priors) via an L2-regularized extension of stratified LD-score regression (S-LDSC)^30^, and subsequently perform fine-mapping using FINEMAP and SuSiE^18^. Briefly, functional annotations included epigenomic and genomic annotations, minor allele frequency (MAF) bins, binary or continuous functional annotations, LD-related annotations such as LD level, predicted allele age, recombination rate, and CpG content^28^. Functionally-informed fine-mapping was also performed using the three LD reference panels.

In total, 16 fine-mapping analyses were conducted (12 multi-variant analyses using four fine-mapping methods and three LD reference panels and four LD-independent single-variant fine-mapping analyses), varying parameters to examine their impact and the convergence of results. “Consensus SNPs” were defined as those in the 95% CS from at least two methods (either statistical and/or functional fine-mapping) that used the same LD option and with a PIP >0.95 or >0.50 (**Table 1**) (24 opportunities for a SNP to be a consensus SNP). The “union consensus” set of SNPs was defined as all consensus SNPs across LD options PIP >0.50, excluding SNPs identified only with the UKB LD reference panel. The numbers of SNPs fine-mapped at PIP >0.95 and PIP >0.50 between different methods and different LD options were compared using two-sided paired t-tests.

All steps of the statistical and functional fine-mapping analyses have been compiled into a high-throughput pipeline named SAFFARI (**S**tatistical **A**nd **F**unctional **F**ine-mapping **A**pplied to GWAS **R**isk Loc**I**). SAFFARI is implemented through Snakemake in a Linux environment^24^, with options to provide sets of GWAS summary statistics, lists of fine-mapping windows, and to specify LD reference panels, in the form of LD matrices or individual-level genetic data (GitHub: https://github.com/mkoromina/SAFFARI).

### Impact of LD options and locus windows on fine-mapping

We aimed to investigate the impact of using an LD reference panel for fine-mapping or performing single variant fine-mapping with no LD, compared with using LD information calculated from the original GWAS data. The latter is typically considered the gold-standard approach, however is difficult in practice due to data availability and sharing restrictions. We performed several comparative analyses and found that the HRC reference panel, a panel that closely resembles the genetic ancestry of the GWAS, achieves comparable fine-mapping resolution with in-sample LD estimates (**Supplemental Note**). We also compared results from fine-mapping the GWS locus windows versus fixed 3Mb windows, which indicated substantial differences between them, and that the GWS locus windows best represent the GWS association signals from the original GWAS (**Supplemental Note**).

### Annotation of union consensus SNPs

Union consensus SNPs were characterized using the Variant Effect Predictor (VEP) (GRCh37) Ensembl release 109^31^. When SNPs were mapped to multiple transcripts, the most severe variant consequence was retained for annotation, and when SNPs fell within intergenic or regulatory regions, no genes were annotated^31^. If annotated genes overlapped and the SNP had the same severity consequence, then both genes were annotated. Additional annotations included the CADD scores (https://cadd.gs.washington.edu/), which denote the likelihood of the variant being deleterious or disease-causing (CADD >= 20) and ClinVar annotations (https://www.ncbi.nlm.nih.gov/clinvar/) describing the association of variants with diseases (i.e., benign, pathogenic, etc). Union consensus SNPs were further annotated with RegulomeDB (v.2.2) to determine whether they have functional consequences and lie in non-coding regions and to annotate them to the relevant regulatory elements^32^. RegulomeDB probability and ranking scores are positively correlated and predict functional variants in regulatory elements. Probability scores closer to 1 and ranking scores below 2 provide increased evidence of a variant to be in a functional region^32^. Probability of being loss-of-function intolerant (pLI) and loss-of-function observed/expected upper bound fraction (LOEUF) scores were retrieved from the Genome Aggregation Database (gnomAD) v4.0.0. Genes were classified as intolerant to loss of function (LoF) variants if LOEUF< 0.6 or pLI ≥0.9. We also used the Open Targets platform^33^ to detect druggable genes amongst our set of high confidence genes for BD risk.

### QTL integrative analyses

Union consensus SNPs were investigated for putative causal relationships with BD via brain gene expression, splicing or methylation, using Summary data-based Mendelian randomization (SMR) (v1.03)^34,35^. Data on expression quantitative trait loci (eQTLs) and splicing quantitative trait loci (sQTLs) were obtained from the BrainMeta study (v2), which comprised RNA-seq data of 2,865 brain cortex samples from 2,443 unrelated individuals of EUR ancestries with genome-wide SNP data^36^. Data on methylation quantitative trait loci (mQTLs) were obtained from the Brain-mMeta study^37^, a meta-analysis of adult cortex or fetal brain samples, comprising 1,160 individuals with methylation levels measured using the Illumina HumanMethylation450K array. We analyzed *cis*-QTLs, which were defined as those within 2 Mb of each gene^36^. Of the union consensus SNPs, 10 were present in the BrainMeta QTL data, and 10 were present in the Brain-mMeta data. Using the BD GWAS^16^ and QTL summary statistics^36^, each union consensus SNP was analyzed as the target SNP for probes within a 2 Mb window on either side using the -- *extract-target-snp-probe* option in SMR. Only probes for which the union consensus SNP was a genome-wide significant QTL (P < 5 × 10^−8^) were analyzed, to ensure robustly associated instruments for the SMR analysis^34,35^. A Bonferroni correction was applied for 13 tests in the eQTL (P^SMR^ < 3.84 × 10^−3^), 57 tests in the sQTL (P_SMR_ < 8.77 × 10^−4^) and 40 tests in the mQTL analyses (P_SMR_ < 1.25 × 10^−3^). The significance threshold for the HEIDI test (heterogeneity in dependent instruments) was *P*_HEIDI_ ≥ 0.0135. The HEIDI test is used to identify potential violations of the Mendelian Randomization assumptions, specifically the assumption of no horizontal pleiotropy. A SNP with passing the Bonferroni-corrected P_SMR_ and the *P*_*HEIDI*_ thresholds indicates either a direct causal role or a pleiotropic effect of the BD-associated SNPs on gene expression, splicing or methylation level.

### Overlap with epigenomic peaks and rare variant association signal

Union consensus SNPs were examined for physical overlap with promoters or enhancers of gene expression in human brain cell-types. Data on epigenomic peaks were obtained from purified bulk, H3K27ac and H3K4me3 ChIP-seq of neurons and astrocytes previously published and used to detect active promoters and enhancers^38^. Physical overlap was visually examined via locus plots using R (R version 4.1.2). For SNPs located in promoters, we assigned the corresponding gene name. For active enhancers, the target gene was assigned based on PLAC-Seq data^38^ on enhancer-promoter interactions. Genes linked to union consensus SNPs via overlap with epigenomic peaks, SMR, or missense annotations, were further assessed for convergence with findings from an exome sequencing study of BD published by the Bipolar Exome (BipEx) Collaboration^39^. Using the BipEx browser^39^, genes annotated to union consensus SNPs were compared for an overlap against BipEx genes characterized by a significant (P < 0.05) burden of either damaging missense or LoF variants.

### Fine-mapping the MHC locus

The major histocompatibility complex (MHC) locus was fine-mapped separately due to its complex genetic variation and long-range LD structure^40^. The human leukocyte antigen (HLA) alleles and amino acid variants were imputed in the PGC BD data, using the 1000 Genomes phase 3 reference panel comprising 503 EUR individuals^41^ with HLA alleles determined via sequencing. This reference was obtained from the CookHLA GitHub repository^42^ (CookHLA v.1.0.1) and included 151 HLA alleles (65 2-digit and 86 4-digit) with a MAF >0.01 and <0.99, 1,213 amino acid variants, and 1,268 SNPs within the MHC region (chromosome 6, 29-34 Mb).

Variation in the MHC was imputed for 48 BD cohorts where individual-level genotyped SNP data were available within the PGC (33,827 BD, 53,953 controls), using IMPUTE2, implemented via the Rapid Imputation and COmputational PIpeLIne for GWAS (RICOPILI)^43^. RICOPILI was used to perform association analysis, under an additive logistic regression model in PLINK v1.90^29^, covarying for the first 5 principal components (PCs) of genetic ancestry and any others associated with case-control status within each cohort, as per the BD GWAS^16^. To control test statistic inflation at variants with low MAF in small cohorts, variants were retained only if cohort MAF was greater than 1% and minor allele count was greater than 10 in either cases or controls (whichever had smaller N). Meta-analysis of the filtered association statistics was conducted using an inverse-variance-weighted fixed-effects model in METAL (version 2011-03-25) via RICOPILI^44^.

Conditional analysis of the MHC-association results was performed to identify whether there are any additional independent associations, by conditioning on the top lead variant within the locus. In brief, the dosage data for the top lead variant in the meta-analysis were extracted for each cohort, converted into a single value representing the dosage of the A1 allele (range 0-2), and this was added as a covariate in the analysis. Association testing, filtering of results per cohort, and the meta-analysis were carried out as described above.

### Polygenic risk scoring

Fine-mapping results were further evaluated by testing whether fine-mapping effect sizes could improve the performance of PRS in independent cohorts using PolyPred^45^, a method which combines effect sizes from fine-mapping with those from a standard PRS approach, such as PRS-CS^46^. PRS were calculated for individuals in 12 testing cohorts of BD cases and controls that were independent of the BD GWAS: three new PGC cohorts of EUR ancestries, two cohorts of East Asian ancestries, four cohorts of admixed-African American ancestries, and three cohorts of Latino ancestries, some of which have been described previously^16^ (**Supplemental Note**).

An analytical workflow outlining the steps of the PolyPred pipeline that we followed is shown in **Supplemental Figure S1**. First, the standard approach used was PRS-CS, which uses a Bayesian regression framework to place continuous shrinkage priors on effect sizes of SNPs in the PRS, adaptive to the strength of their association signal in the BD GWAS^16^, and the LD structure from an external reference panel^46^. The UKB EUR ancestry reference panel was used to estimate LD between SNPs, matching the ancestry of the discovery GWAS^16^. PRS-CS yielded weights for approximately 1 million SNPs to be included in the PRS. Second, genome-wide fine-mapping was performed on the BD GWAS summary statistics^16^, using both SuSiE and PolyFun-SuSie as previously described, with LD information obtained from the HRC reference panel, to derive causal effect sizes for all SNPs across the genome. Third, PolyPred was used to combine the SNP weights from PRS-CS with SuSie effect sizes (“SuSie+PRS-CS”) and SNP weights from PRS-CS with PolyFun-SuSiE effect sizes (“Polypred-P”). Briefly, Polypred “mixes” the effect sizes from the two predictors via the non-negative least squares method, assigning a weight to each predictor that yields the optimally performing PRS in a specific testing cohort. Each testing cohort was used to tune the optimal PolyPred weights. Fourth, three PRS were calculated for each individual in the testing cohorts, using PLINK v1.90^29^ to weight SNPs by their effect sizes from PRS-CS, SuSiE+PRS-CS and Polypred-P respectively, and sum across all SNPs in each PRS. Finally, PRS were tested for association with case versus control status in each testing cohort using a logistic regression model including PCs as necessary to control for genetic ancestry^47^. In each testing cohort, the amount of phenotypic variance explained by the PRS (R^2^) and the 95% confidence intervals were calculated on the liability scale^48^, using the r2redux R package^49^, assuming a lifetime prevalence of BD in the general population of 2%. The R^2^ of each fine-mapping-informed PRS was statistically compared against the R^2^ of PRS-CS using the r2redux package (r2_diff function)^49^. In addition, we computed the effective sample size-weighted combined R^2^ values from PRS across different ancestries. Specifically, we transformed each R^2^ to a correlation coefficient, applied the Fisher Z transformation, computed the effective sample size (Neff)-weighted mean of the Fisher Z values, and then back-transformed to obtain a combined R^2^.

## Results

### Fine-mapping identifies likely causal BD variants

Stepwise conditional analyses using GCTA-COJO were performed in each of the 64 BD GWS loci (**Supplemental Table S2**), conditioning associations on their top lead SNP and any subsequent conditionally independent associations, to identify loci that contained independent signals (conditional *P*⍰<⍰5×10^−6^). This analysis supported the existence of one association signal at 62 loci (**Supplemental Table S3**), and two independent association signals within the *MSRA* locus on chromosome 8 and the *RP1-84O15*.*2* locus on chromosome 8 (**Supplemental Table S3**).

Excluding the MHC, GWS loci were fine-mapped via a suite of Bayesian fine-mapping tools (SuSiE, FINEMAP, PolyFun+SuSiE, PolyFun+FINEMAP) to prioritize SNPs likely to be causal for BD, and examine the impact of different LD reference options (see Methods). **Figure 2** shows the number of SNPs with a PIP >0.95 and PIP >0.50 in each fine-mapping analysis, alongside the Jaccard Index of concordance between each pair of the 16 fine-mapping analyses, calculated based on SNPs with PIP > 0.5 and part of a 95% credible set. A breakdown of the Jaccard Indices for analyses grouped by LD option, statistical or functional fine-mapping and fine-mapping method are provided in the **Supplemental Figure S2**.

**Figure 2.**
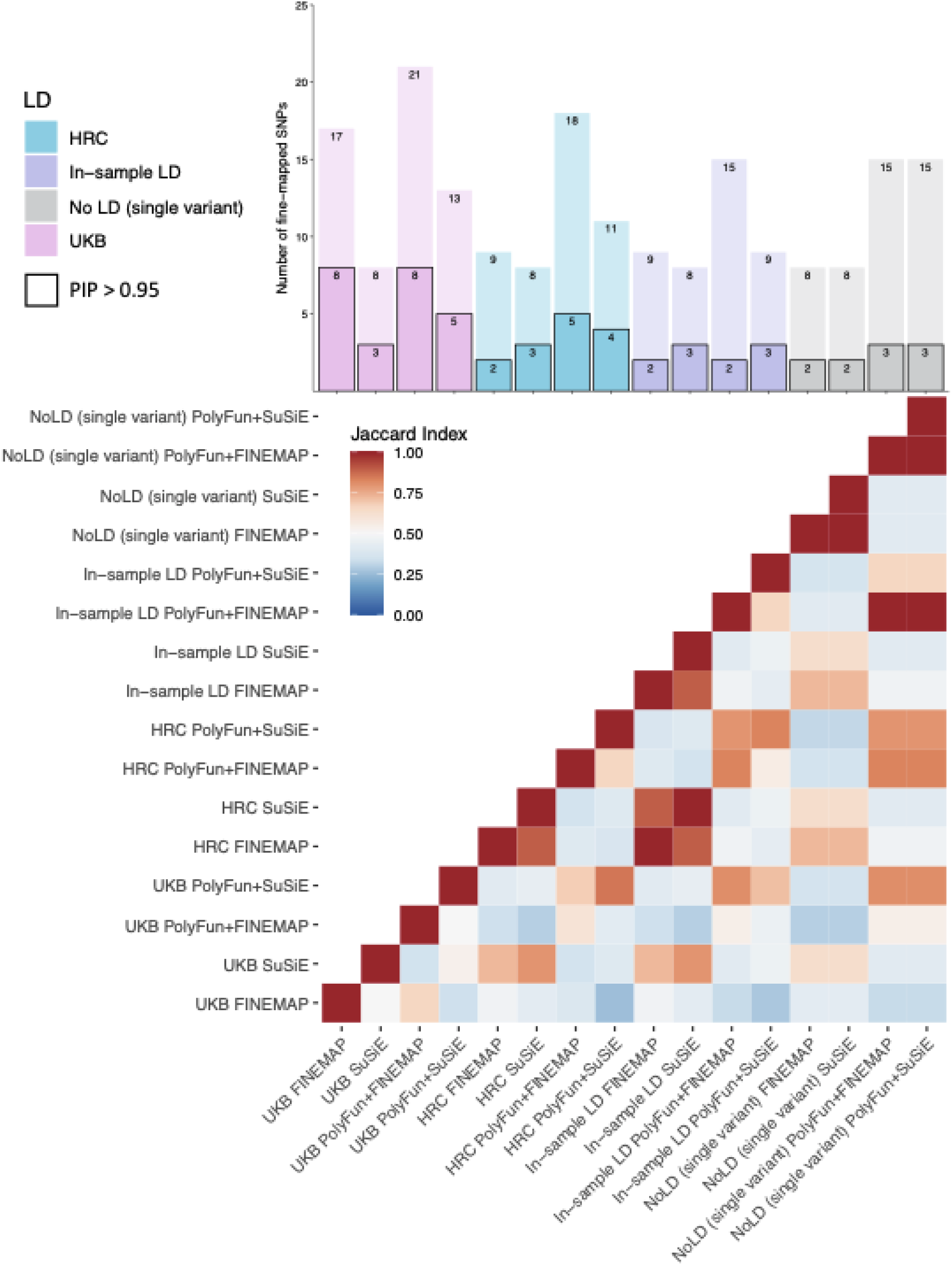
Results and comparison of 16 fine-mapping analyses conducted. The barplot displays the number of SNPs fine-mapped with PIP > 0.5 and part of a 95% credible set on the y-axis and each fine-mapping analysis on the x-axis. The black bordered bars indicate the number of SNPs fine-mapped with PIP > 0.95 and part of a 95% credible set. Each analysis is named according to [LD option]_[fine-mapping method]. The heatmap displays the Jaccard Index of concordance between each pair of fine-mapping analyses, calculated based on SNPs with PIP > 0.5 and part of a 95% credible set. Jaccard Indices ranged from 0.25 and 1, with a mean Jaccard Index of 0.54 (SD = 0.20).

Functional fine-mapping analyses yielded significantly more fine-mapped SNPs compared with the corresponding statistical fine-mapping analyses at PIP > 0.95 and PIP > 0.5 (P = 6.47×10^−4^ and P = 0.03 respectively) (**Figure 2**). There were no significant differences in the numbers of SNPs fine-mapped between the four LD options, between the two statistical fine-mapping methods or between the two functional fine-mapping methods. Approximately a quarter of GWS loci (N= 15) had high PIP SNPs (>0.50). Employing different fine-mapping methods and LD reference panels revealed a substantial number of consensus SNPs with PIP >0.50 (17 SNPs), but fewer met the stricter threshold of PIP >0.95 (6 SNPs) (**Figure 3**). The number of 95% credible sets per locus varied based on the fine-mapping method (**Supplemental Figure S3**).

**Figure 3.**
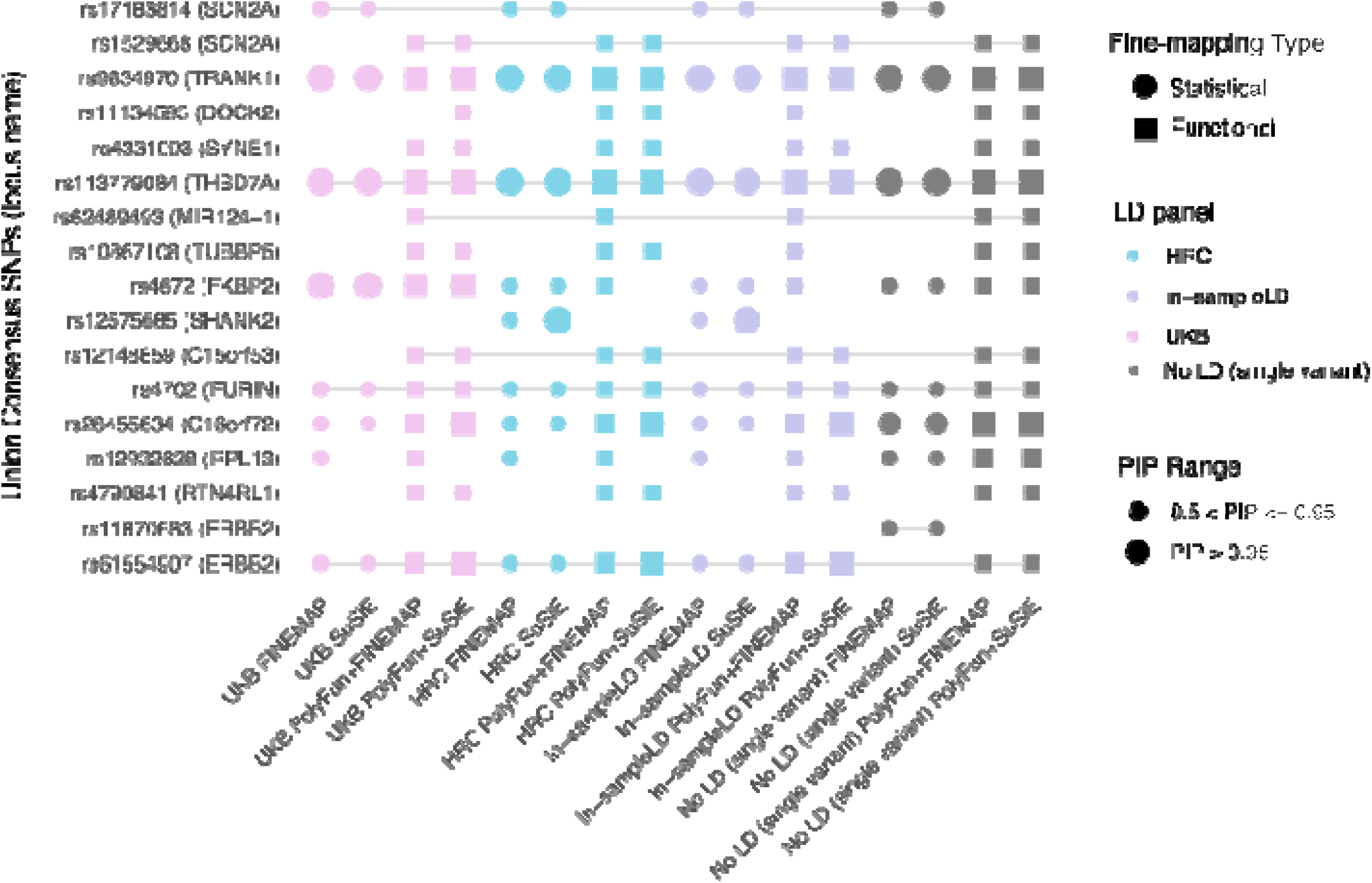
Plot of union consensus SNPs across all 16 fine-mapping analyses, including different LD options and fine-mapping methods. The color of the points corresponds to the LD option used: UK Biobank (pink), Haplotype Reference Consortium (blue), in-sample LD (purple) and no LD (single variant fine-mapping) (grey). Circles indicate statistical fine-mapping methods and squares indicate functional fine-mapping methods. Small shapes denote SNPs with PIP between 0.50 and 0.90, while large shapes denote SNPs with PIP above 0.95. On the y axis, the genome-wide significant locus names, as defined in the origins GWAS paper, are in parenthesis after each SNP. On the x-axis, analyses are named according to [LD option] [fine-mapping method].

The smallest 95% CS per locus for every fine-mapping method and LD reference panel (**Supplemental Figure S3**) was also calculated. Approximately ⅕ (N= 10-19) or ½ (N= 32-41) of the 63 fine-mapped loci had 95% CSs with a small number of SNPs (N_SNPs_< 10). The percentage of fine-mapped loci harboring 95% CSs with N_SNPs_< 10 was dependent on the fine-mapping method, with FINEMAP and PolyFun+FINEMAP harboring smaller 95% CSs and SuSiE and PolyFun+SuSiE larger 95% CSs.

The union consensus set (PIP >0.5) comprised 17 SNPs (from 15 GWS loci), indicating that many of the same SNPs were prioritized regardless of which LD reference panel was used (**Figure 3**). There were 15 SNPs consistently prioritized as the likely causal variant across all LD options (**Figure 3, Supplemental Figure S4**). The distribution of SNPs with PIP >0.50 for each GWS locus across different methods and LD options is provided in the **Supplemental Figure S4**.

Variant annotation of the union consensus SNPs via VEP^31^ indicated that 5 of the 17 fall in intronic regions (**Supplemental Table S4**). Two of the union consensus SNPs are missense variants: rs17183814 in *SCN2A* (CADD: 20, ClinVar benign for seizures and developmental and epileptic encephalopathy) and rs4672 in *FKBP2* (CADD: 22.5, not in ClinVar). More details about the variant annotations of the union consensus SNPs through different online databases is provided in **Supplemental Table S4**.

### QTL integrative analyses and overlap with epigenomic peaks

Summary data-based Mendelian randomization (SMR)^34,35^ was used to identify putative causal relationships between union consensus SNPs and BD via gene expression, splicing or methylation, by integrating the BD GWAS association statistics with brain eQTL, sQTL and mQTL summary statistics. eQTL and sQTL data were based on the BrainMeta study (2,865 brain cortex samples from 2,443 unrelated individuals of EUR ancestries)^36^ and mQTL data were from the Brain-mMeta study (adult cortex or fetal brain samples in 1,160 individuals)^37^. Union consensu SNPs with genome-wide significant cis-QTL P values (P < 5×10^−8^) and their corresponding gene expression, slicing or methylation probes were selected as target SNP-probe pairs for SMR, yielding 13, 57 and 40 SNP-probe pairs for eQTL, sQTL and mQTL analyses respectively. In the eQTL analyses, there were 5 union consensus SNPs with significant P_SMR_ that passed the HEIDI (heterogeneity in dependent instruments) test for 9 different genes, suggesting that their effect on BD is mediated via gene expression in the brain (**Figure 4, Supplemental Table S5**). Three of the union consensus SNPs showed evidence of causal effects on BD via expression of more than one gene in their cis-region. In the sQTL analyses, there were 6 union consensus SNPs with significant P_SMR_ results, and passing the HEIDI test, implicating 11 genes (**Figure 4, Supplemental Table S5**). In the mQTL analyses, there were 20 SNP-probe pairs passing the P_SMR_ and P_HEIDI_ thresholds; of which two methylation probes were annotated to specific genes (*FKBP2* and *PLCB3*) (**Figure 4, Supplemental Table S5**).

**Figure 4.**
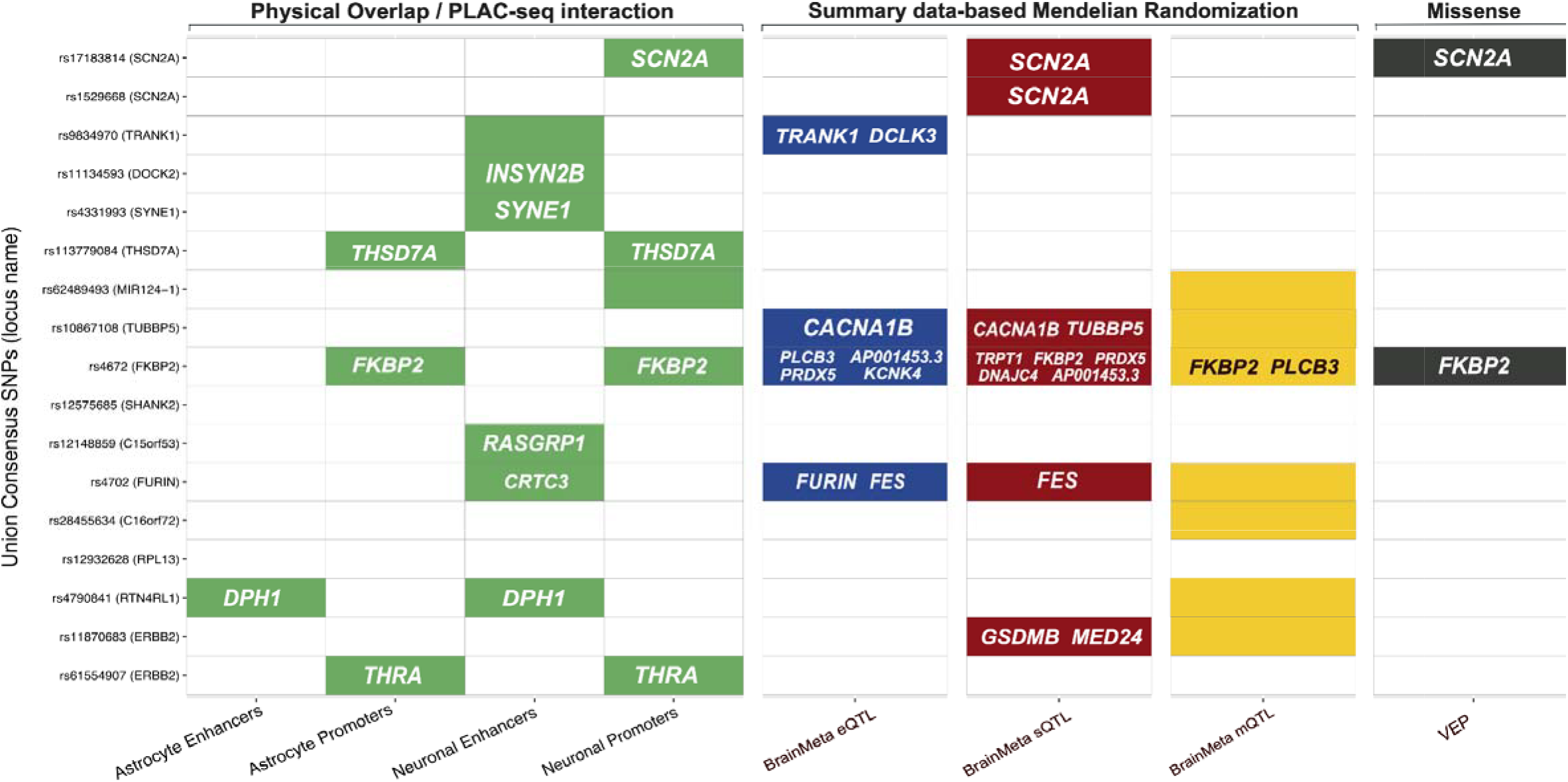
Summary of analyses performed to link each fine-mapped SNP to the relevant gene(s). The y-axis shows the 17 union consensus SNPs and the name of the corresponding genome-wide significant locus (as defined in the original GWAS). On the x-axis, the columns depict the results of 8 analyses performed to link the fine-mapped SNPs to the relevant gene(s). The analysis method and the dataset used are labeled above and below the figure respectively. Colored cells denote significant results and the relevant gene names are printed within each cell. For fine-mapped SNPs located in active enhancers, the relevant genes were obtained using data on PLAC-seq interactions with gene promoters. A colored cell includes no gene name when there was no known interaction between the enhancer and a promoter, or when the methylation probe was not annotated to any gene. Empty cells are those with non-significant results, or where the SNP was not present in the dataset used.

There were 11 union consensus SNPs that physically overlapped with active enhancers or promoters of gene expression in brain cell-types^38^, particularly neurons (**Figure 4**). Four union consensus SNPs were located in active promoters of the *SCN2A, THSD7A, FKBP2* and *THRA* genes. Through the utilization of PLAC-seq data, we explored enhancer-promoter interactions, specifically for enhancers in which there is a physical overlap with the union consensus SNPs, and prioritized their genes (**Figure 4**). Amongst the implicated target genes through enhancer-promoter interactions are *INSYN2B, SYNE1, RASGRP1, CRTC3, DPH1* and *THRA*.

### Candidate risk genes based on convergence of evidence

By aggregating multiple lines of fine-mapping validation evidence, we present results for high-confidence genes for BD. Specifically, a gene was characterized as high-confidence if it was linked to a fine-mapped SNP via active promoters or enhancers, brain gene expression, splicing or methylation, or if the fine-mapped SNP was a missense variant (**Figure 4, Supplemental Figure S5**). Assuming that a single variant may act through multiple risk genes, we took the union of the prioritized genes across the different lines of evidence described above. Taken together, the data support the roles of the following 23 genes in BD: *SCN2A, TRANK1, DCLK3, INSYN2B, SYNE1, THSD7A, CACNA1B, TUBBP5, PLCB3, AP001453*.*3, PRDX5, KCNK4, CRTC3, TRPT1, FKBP2, DNAJC4, RASGRP1, FURIN, FES, DPH1, GSDMB, MED24* and *THRA* (**Supplemental Table S6**). **Supplemental Figure S5** provides multi-track locus plots depicting GWAS association statistics, fine-mapping results, overlap with epigenomic peaks from neurons or astrocytes and gene tracks for the majority of GWS loci. We assessed the high-confidence genes for evidence of rare variant associations with BD, using data from the BipEx exome sequencing study^39^. Amongst the 23 genes examined, *THSD7A, CACNA1B, SCN2A* and *TRANK1* had a significant burden (*p* < 0.05) of damaging missense or LoF variants in BD versus controls. Many high-confidence genes were classified as druggable based on Open Targets platform (*SCN2A, CACNA1B, PRDX5, THRA, MED24, SYNE1, KCNK4, FKBP2, RASGRP1, PLCB3, DCLK3, FURIN, FES*). Detailed literature information about the biological relevance of the high-confidence genes can be found in the **Supplemental Table S6**.

### Dissecting the MHC locus

In the original GWAS, the most significant SNP in the extended MHC was rs13195402 (26.4 Mb, P = 5.8×10^−15^) which is a missense variant in *BTN2A1*. Conditional analysis on this SNP suggested a single association signal across the extended MHC, and there were no associations between structural haplotypes of the complement component 4 genes (*C4A*/*C4B*) (∼31.9 Mb) and BD^16^. Here, we performed association analyses of variants in the MHC region (chromosome 6, 29-34 Mb) including *HLA* alleles, amino acids, SNPs and insertion/ deletion variants, in a sample of 33,781 BD cases and 53,869 controls. The most significant variant in the classical MHC was rs1541269 (30.1 Mb, P = 6.71×10^−12^, LD r^2^ = 0.55 with the original index SNP rs13195402 in European populations)^16^. While initially some variants in *HLA* genes reached GWS (**Supplemental Table S7**), none remained after conditioning on rs1541269, suggesting the associations were driven by LD with more strongly associated variants located upstream (**Supplemental Figure 6, Supplemental Table S8**).

### Leveraging fine-mapping to improve BD polygenic risk scores

We assessed whether fine-mapping results could be used to improve the performance of BD PRS in 12 testing cohorts: three EUR cohorts that were independent of the BD GWAS, two East Asian cohorts, four admixed African American cohorts, and three Latino cohorts^47,50,51^. Standard PRS were calculated using the PRS-CS method, and fine-mapping informed PRS were calculated via PolyPred, to integrate statistical fine-mapping results (SuSiE+PRS-CS) or functional fine-mapping results (Polypred-P). Across PRS methods, PRS were significantly higher in BD cases versus controls in all EUR target cohorts and most non-EUR cohorts (**Figure 5, Supplemental Tables S9**). Using PRS-CS, the effective sample size-weighted phenotypic variance explained on the liability scale was 12.26% in EUR ancestries, 2.41% in East Asian ancestries, 0.20% in African American ancestries and 0.28% in Latino ancestries (**Figure 5, Supplemental Table S10**). Examining fine-mapping-informed PRS, SuSiE+PRS-CS or Polypred-P explained more phenotypic variance than PRS-CS in all cohorts, with PolyPred-P showing the best performance (**Figure 5**). However, increased variance explained by SuSiE+PRS-CS or Polypred-P compared with PRS-CS, was only statistically significant in the Japanese BD cohort (P = 1.22×10^−5^ and P = 2.29×10^−6^ respectively), one African American (P = 0.035 and P = 0.044 respectively) and one Latino cohort (P = 0.046 and P = 0.002 respectively) (**Supplemental Table S9, Figure 5**).

**Figure 5.**
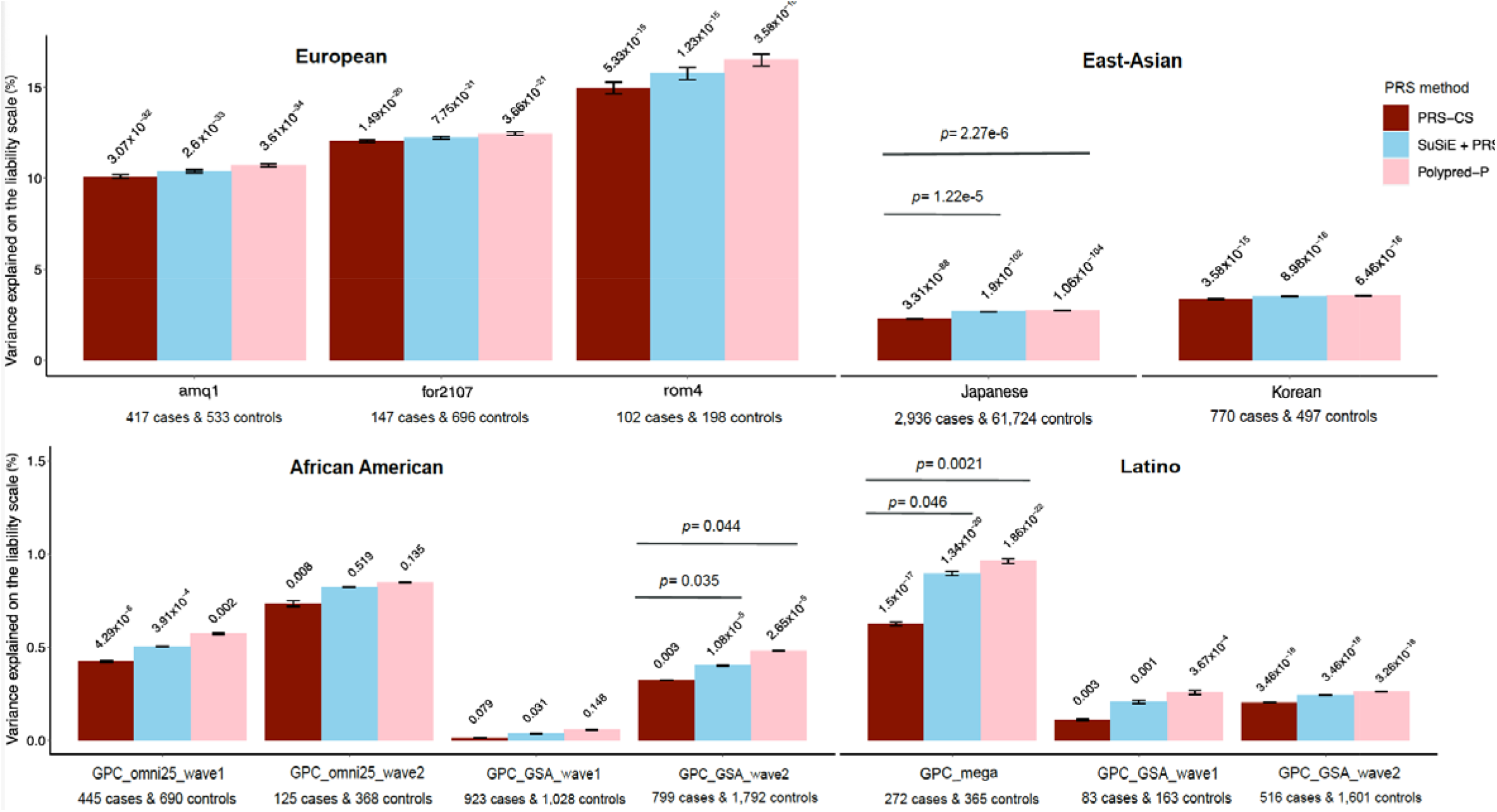
Phenotypic variance in BD explained by standard PRS (PRS-CS) and fine-mapping-informed PRS (SuSiE+PRS-CS and PolyPred-P) in target cohorts of diverse genetic ancestries. The x-axis displays the target cohorts, grouped by genetic ancestry, and the PRS method used. The name of each cohort and the number of BD cases and controls is shown below each barplot. The y-axis shows the percentage variance explained on the liability scale, assuming a 2% population prevalence of BD. Error bars represent 95% confidence intervals on the variance explained. P-values for the association of PRS with case versus control status are printed on top of each bar. Significant P-values (P < 0.05) for the test of difference in variance explained by the fine-mapping informed PRS versus PRS-CS are provided above the horizontal lines.

## Discussion

In the most comprehensive fine-mapping study of BD GWAS risk loci to date, we applied a suite of statistical and functional fine-mapping methods to prioritize 17 likely causal SNPs for BD in 15 genomic loci. We linked these SNPs to genes and investigated their likely functional consequences, by integrating variant annotations, brain cell-type epigenomic annotations and brain QTLs. Convergence of evidence across these analyses prioritized 23 high-confidence genes, which are strong candidates for functional validation experiments to understand the mechanisms by which they increase risk of BD.

We defined a union consensus set of SNPs representing those likely causal for BD based on the convergence between fine-mapping methods and LD reference panels. This comprised 17 SNPs (from 15 GWS loci), indicating that many of the same SNPs were prioritized across fine-mapping analyses (**Figure 3**). Linking these SNPs to genes and investigating their likely functional consequences using computational approaches and relevant datasets, prioritized 23 high-confidence genes (**Figure 4**). Overall, we hypothesized that a single putative causal SNP may influence multiple genes due to various factors such as the impact of enhancer elements on multiple genes’ expression, overlap of eQTLs and sQTLs with epigenomic annotations and missense variants, and overlapping genomic coordinates of genes^36,52,53^.

This study uncovered novel insights into BD. Six of the genes prioritized have synaptic functions, including two with presynaptic and four with postsynaptic annotations. The functions of these genes encompass both cellular excitability (regulation of neurotransmitter levels and membrane potential) and cellular organization (arrangement of the actin cytoskeleton, endocytosis, and the postsynaptic specialization). Prioritized genes implicate a variety of neurotransmitters, both excitatory and inhibitory. These findings highlight the impact of BD risk variants on diverse aspects of synaptic signaling. While all prioritized genes are expressed in the brain and most display enrichment of expression in several brain cell types, three of the genes prioritized have enhanced expression in cells of the gut, including gastric mucous secreting cells, and proximal and distal enterocytes. These cells play roles in intestinal permeability, inflammation and the enteric nervous system, and our findings lend genetic support to the involvement of the microbiota–gut–brain axis in BD^65^. The *PLCB3, KCNK4*, and *DPH1* genes prioritized have previously been linked to neurodevelopmental delay^58–60^ but not BD. Our study also provides novel insights into the potential molecular mechanisms underlying known BD risk genes. For example, results suggest that fine-mapped variants impact BD through alternate splicing of *SCN2A* and *CACNA1B* in the brain, findings which may inform functional laboratory experiments.

In the MHC, there were several polymorphic alleles and amino acid variants in the *HLA-C* and *HLA-B* genes associated with BD at GWS (chromosome 6, 31.2-31.3 Mb). The *HLA-C*07:01* and *HLA-B*08:01* alleles were negatively associated with BD, in line with previous studies reporting their protective effects on SCZ^66,67^. However, these associations were removed after conditioning on the top lead variant in the MHC (rs1541269, 30 Mb), suggesting the effects were driven by LD with more strongly associated variants located upstream. This is consistent with published findings in the PGC BD data, showing no association between the structural variants in the complement component 4 genes (C4A/C4B) (∼31.9 Mb) and BD, either before or after conditioning on the most associated MHC SNP (rs13195402, 26.4 Mb)^16^. Overall, this analysis of HLA variation in BD again suggests a single association signal across the MHC, and that the causal variants and genes are outside the classical MHC locus, in contrast to findings in schizophrenia^68^.

Fine-mapping-informed PRS, developed by combining GWAS effect sizes and genome-wide fine-mapping effect sizes using PolyPred, explained a greater proportion of phenotypic variance, compared with PRS based on GWAS effect sizes alone. This adds support to our fine-mapping results, as leveraging information on causal effect sizes rather than relying solely on association statistics should improve genetic risk prediction. Under the assumption that the causal variants are shared across ancestries, we anticipated that fine-mapping-informed PRS would improve the transferability of BD PRS into diverse genetic ancestries. Indeed, there was a modest increase in the phenotypic variance explained relative to standard PRS in all genetic ancestry groups. However, the performance of all PRS in non-European cohorts still lagged greatly behind that in Europeans (**Figure 5, Supplemental Table S9, S10**), emphasizing the need for larger studies in diverse genetic ancestries and further development of methods to improve PRS transferability between ancestries.

Our strategy of applying a suite of fine-mapping methods and examining the convergence of the results was driven by the variety of the underlying fine-mapping algorithms, and their corresponding strengths and limitations. Consistent with previous literature, we detected more SNPs with high PIPs when incorporating functional priors using PolyFun^18^. As expected, the specification of LD structure, fine-mapping window, and number of causal variants impacted fine-mapping results. Considering “in-sample” LD from the PGC BD data (albeit a subset of cohorts that were available) as the gold-standard, using the HRC reference panel yielded the most similar fine-mapping results. This observation may be explained by the HRC being used as an imputation reference panel for almost all cohorts in the GWAS (53/ 57 cohorts). Results suggest that a large and well-matched LD reference panel to the GWAS sample can be used to achieve high-quality fine-mapping results. This has advantageous implications in scenarios when calculating in-sample LD is not possible due to data sharing restrictions, or when obtaining LD information from many cohorts becomes increasingly challenging as GWAS meta-analyses grow. Moreover, although conditional analysis indicated one causal variant per GWS locus, our results are highly consistent when using LD reference panels and allowing up to 5 causal variants per GWS locus. The latter analyses also yielded a greater number of likely causal SNPs. To facilitate rapid and scalable fine-mapping of GWAS loci, we developed a fine-mapping pipeline (GitHub: https://github.com/mkoromina/SAFFARI) with options to specify multiple fine-mapping methods, GWAS summary statistics, fine-mapping windows, and LD reference panels.

Several limitations of this study and future directions must be noted. First, our fine-mapping focused exclusively on EUR ancestry data, owing to the composition of the PGC BD GWAS. However, this enabled us to investigate the impact of LD reference panels on fine-mapping, which would be challenging for diverse ancestry data, given the limited availability of such panels at present. Increasing ancestral diversity in BD GWAS is an active area of research^47^ and in future, the differences in LD structure between populations could be leveraged to aid fine-mapping^69^ and PRS predictions^45^. Second, we approximated “in-sample LD” of the GWAS as we only had access to a subset of the individual-level data (73% of the total effective sample size), we used best guess genotypes to represent imputed dosages, and we merged genotypes across cohorts and calculated LD, in contrast to the GWAS, which was a meta-analysis between cohorts. Third, we applied a conservative approach focusing on SNPs with high PIPs (>0.50), that were part of credible sets, and were supported by different fine-mapping methods. Thus, we prioritized likely causal variants or genes at 15 of the 64 GWS loci. The improvements in PRS performance after integrating genome-wide fine-mapping results, suggest that our analyses capture meaningful information on causality in other genomic regions that did not meet the stringent criteria we applied to fine-map GWS loci. Fourth, these statistical analyses prioritize variants and genes with high-probabilities of being causal risk factors for BD, however computational approaches fall short of proving causality, and have limited capacity to uncover mechanisms. Finally, the enhancer, promoter and QTL data used may be incomplete due to cell-type or context-specific effects, or incomplete mapping of active enhancers to their target genes, and therefore some union consensus SNP effects may not have been detected in our analysis.

In summary, we conducted a comprehensive statistical and functional fine-mapping analysis of BD genomic loci, yielding a resource of likely causal genes and variants for the disorder. These genes and variants now require investigation in functional laboratory experiments to validate their roles, understand mechanisms of risk, and examine opportunities for therapeutic intervention in BD.

## Data Availability

Description of data availability can be found in the corresponding section of the manuscript.

## Data availability

The PGC’s policy is to make genome-wide summary results public. Genome-wide fine-mapping results will be made available through the PGC website upon publication (https://www.med.unc.edu/pgc/results-and-downloads). Individual-level genetic data are accessible via Secondary Analysis Proposals to the Bipolar Disorder Working Group of the PGC (https://www.med.unc.edu/pgc/shared-methods/how-to/). This study included some publicly available datasets accessed through dbGaP - PGC bundle phs001254.v1.p1.

## Code availability

Analysis scripts are available online at [Github: https://github.com/mkoromina/SAFFARI]. All software used is publicly available at the URLs or references cited.

## Acknowledgements

For the purposes of open access, the author has applied a Creative Commons Attribution (CC BY) license to any Accepted Author Manuscript version arising from this submission. We thank the participants who donated their time, life experiences and DNA to this research and the clinical and scientific teams that worked with them. This project was funded by the Baszucki Brain Research Fund via the Milken Institute Center for Strategic Philanthropy. We are deeply indebted to the investigators who comprise the PGC. The PGC has received major funding from the US National Institute of Mental Health (PGC4: R01MH124839, PGC3: U01 MH109528; PGC2: U01 MH094421; PGC1: U01 MH085520). Statistical analyses were carried out on the NL Genetic Cluster Computer (http://www.geneticcluster.org) hosted by SURFsara and the Mount Sinai high performance computing cluster (http://hpc.mssm.edu), which is supported by the Office of Research Infrastructure of the National Institutes of Health under award numbers S10OD018522 and S10OD026880. The content is solely the responsibility of the authors and does not necessarily represent the official views of the National Institutes of Health. Full acknowledgements are included in the Supplemental Note. JRIC is supported by a grant from the Medical Research Foundation (MRF-001-0012-RG-COLE-C0930) and by the NIHR Maudsley Biomedical Research Centre at South London and Maudsley NHS Foundation Trust and King’s College London. The views expressed are those of the authors and not necessarily those of the NIHR or the UK Department of Health and Social Care. EV thanks the support by CIBER - Consorcio Centro de Investigación Biomédica en Red-(CB07/09/0004), Instituto de Salud Carlos III, Spanish Ministry of Science and Innovation and grants PI18/00805 and PI21/00787, integrated into the Plan Nacional de I+D+I and co-financed by the ISCIII-Subdirección General de Evaluación and the Fondo Europeo de Desarrollo Regional (FEDER); the Instituto de Salud Carlos III; the Secretaria d’Universitats i Recerca del Departament d’Economia i Coneixement (2021 SGR 01358), the CERCA Programme, and the Departament de Salut de la Generalitat de Catalunya for the PERIS grant SLT006/17/00357. Thanks also for the support of the European Union Horizon 2020 research and innovation program (EU.3.1.1. Understanding health, wellbeing and disease: Grant No 754907 and EU.3.1.3. Treating and managing disease: Grant No 945151). PBM has been funded through an Australian National Health and Medical Research Council Investigator Grant (1177991). Work for the Japanese cohort was supported by Japan Agency for Medical Research and Development (AMED) grants 22wm0425008, 21ek0109555, 21tm0424220, 21ck0106642, 23ek0410114 and 23tm0424225, Japan Society for the Promotion of Science (JSPS) KAKENHI grant 21H02854 and JP20H00462. Work for the ‘for 2107’ cohort was supported through funds by the German Research Foundation (DFG grants FOR2107 KI588/14-1, and KI588/14-2, and KI588/20-1, KI588/22-1 to Tilo Kircher, Marburg, Germany). Biosamples and corresponding data were sampled, processed and stored in the Marburg Biobank CBBMR. Ethics approval was obtained from the ethics committees of the Medical Schools of the Universities of Marburg (approval identifier Studie 07/2014) and Münster, respectively, in accordance with the Declaration of Helsinki. All subjects volunteered to participate in the study and provided written informed consent.

