## Supplemental Note for "Fine-mapping genomic loci refines bipolar disorder risk genes"

**Contents**

**Supplementary Figures	2**

**Supplementary Note	26**

**Comparison of the impact of LD reference panels on fine-mapping	26**

**Comparison of the impact of locus windows on fine-mapping	28**

**Description of testing cohorts used for polygenic risk scoring analyses	30**

**Full Acknowledgments	33**

**Funding sources	38**

**References	44**

### **Supplementary Figures**

**Figure S1.** **Workflow for computing standard and fine-mapping-informed polygenic risk scores (PRS).** The base GWAS is the published bipolar disorder (BD) GWAS by the Psychiatric Genomics Consortium (PGC). There were 12 target cohorts of BD cases and controls that were independent of the BD GWAS: three new PGC cohorts of EUR ancestries, two cohorts of East Asian ancestries, four cohorts of admixed-African American ancestries, and three cohorts of Latino ancestries.

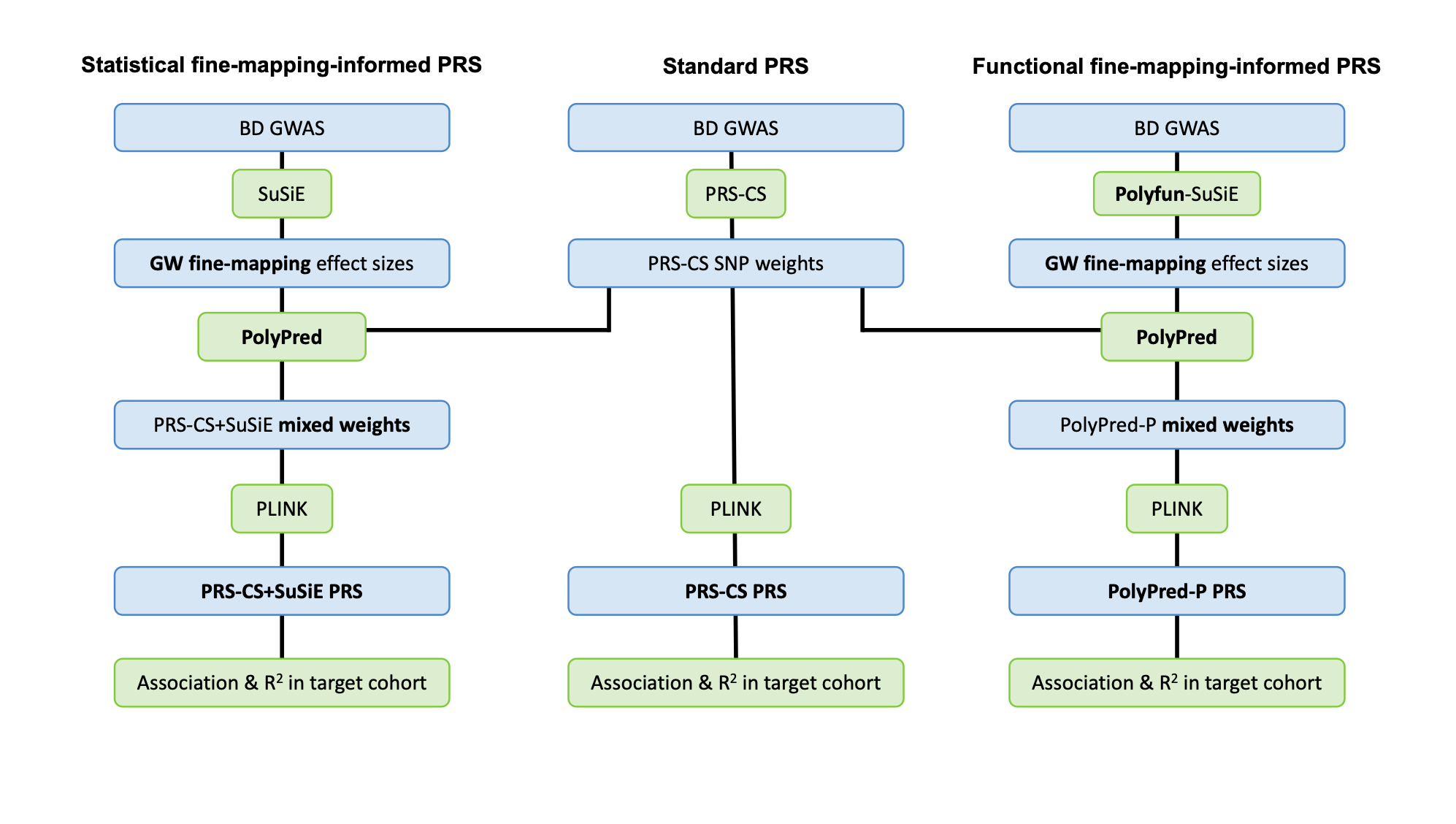

**Figure S2 A-C Heatmaps of Jaccard indices for analyses grouped by A) LD option, B) statistical or functional fine-mapping and C) fine-mapping method.** Jaccard Indices are calculated based on fine-mapped SNPs with PIP > 0.50 and part of 95% credible sets. The mean Jaccard Index and standard deviation (excluding the diagonal elements) is shown above each heatmap. Analyses are named according to [LD option] [fine-mapping method].

**A) Jaccard Indices for analyses grouped by LD option**

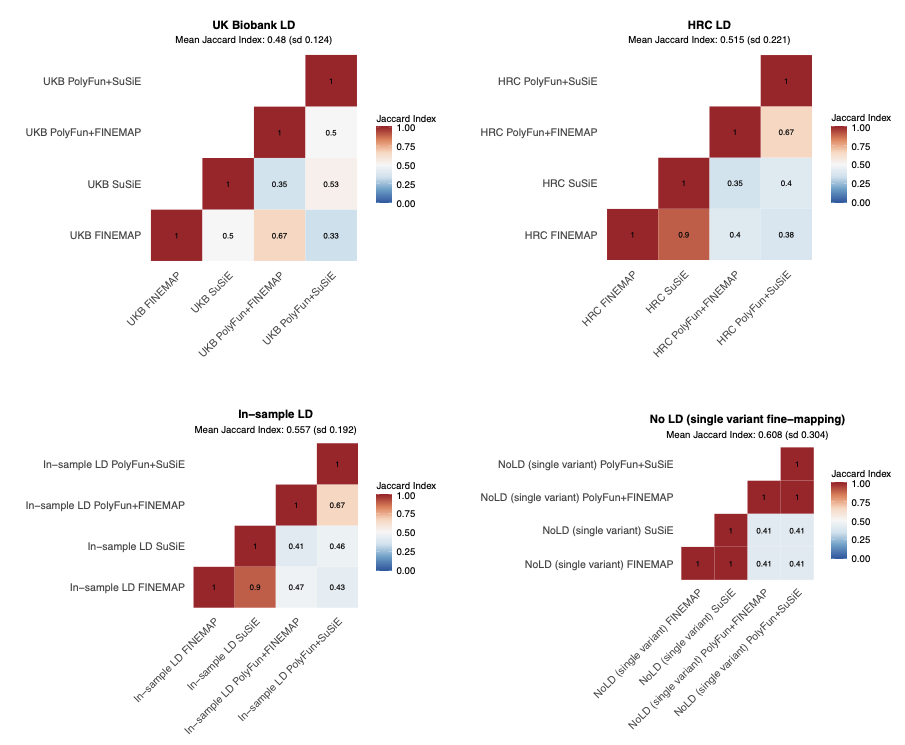

**B) Jaccard Indices for analyses grouped by statistical and functional methods**

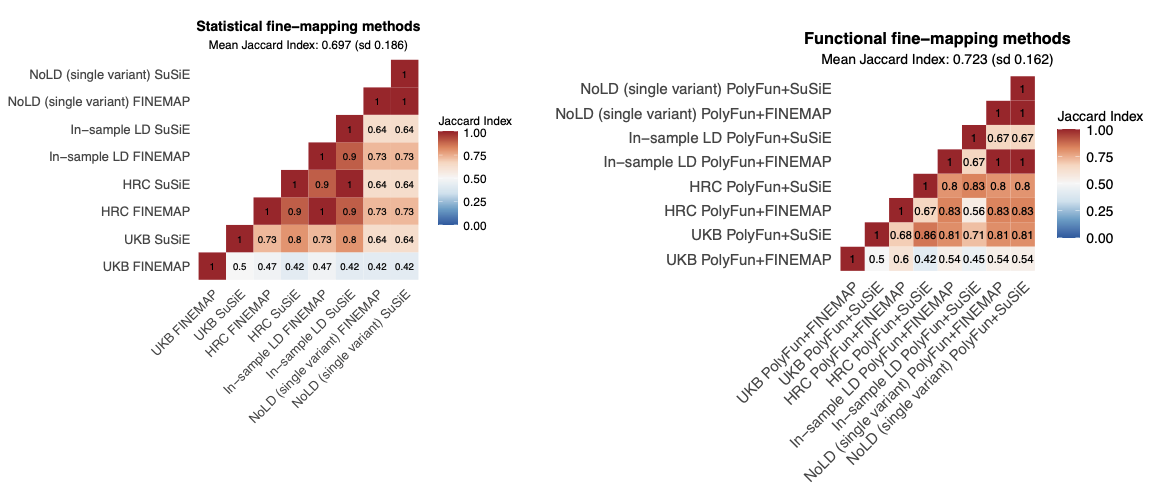

**C) Jaccard Indices for analyses grouped by fine-mapping method**

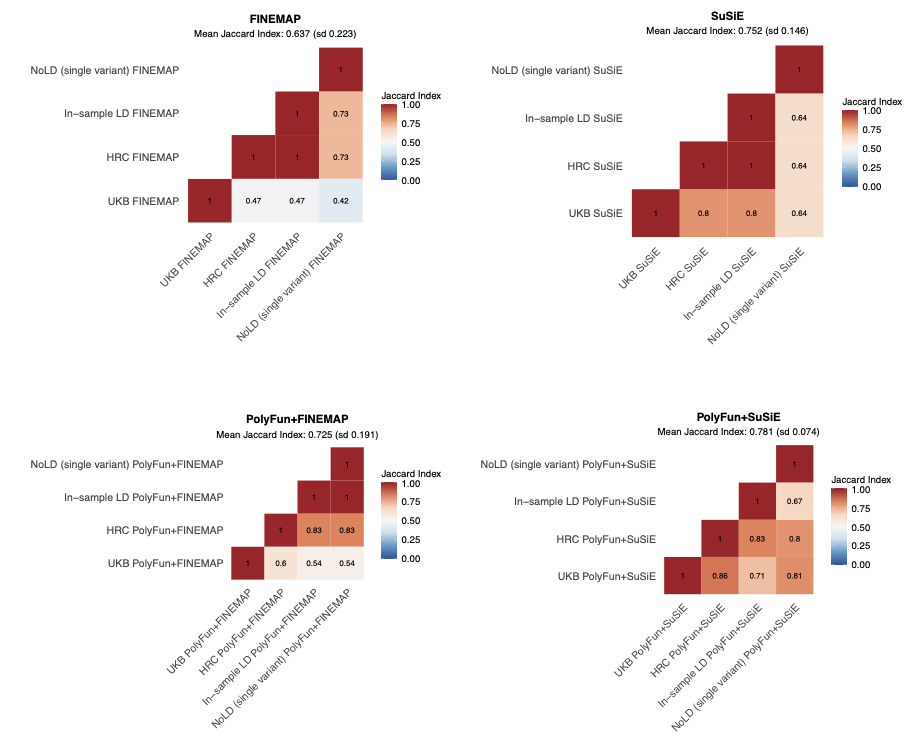

**Figure S3 A-C** **Distribution of fine-mapped loci according to the smallest 95% credible set (95% CS) formed, using different fine-mapping methods and LD reference panels.** Fine-mapping methods include: FINEMAP, SuSiE, PolyFun + FINEMAP, PolyFun + SuSiE. ‘Smallest CS size’ denotes the number of SNPs comprising the smallest 95% CS for a given fine-mapped locus. Absolute numbers denote the number of fine-mapped loci in each ‘smallest CS size’ category. If the ‘smallest CS size’ is 0, this denotes that no 95% CS was formed. Note: 95% CS for FINEMAP is a set of SNPs of which the joint posterior probability of including the causal SNP(s) is higher than 0.95. 95% CS for SuSiE denotes the sum of the PIPs equals to 95%, in which case each PIP is a marginal posterior probability for a single SNP. HRC - Haplotype Reference Consortium, UKB - UK Biobank.

*A). HRC LD reference panel*

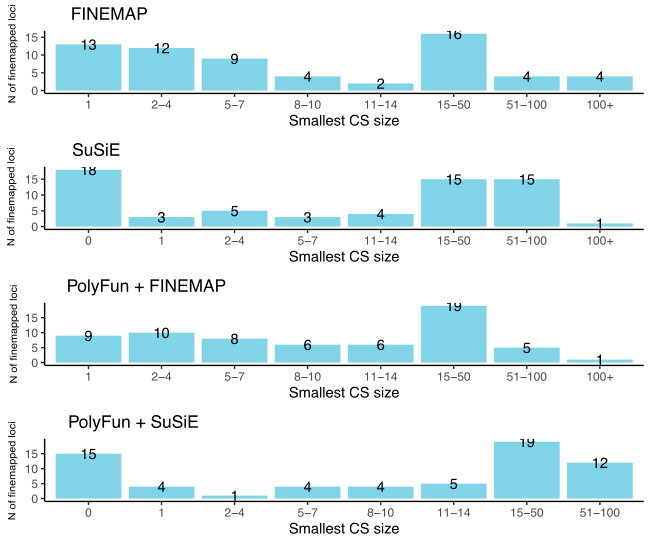

*B). UKB LD reference panel*

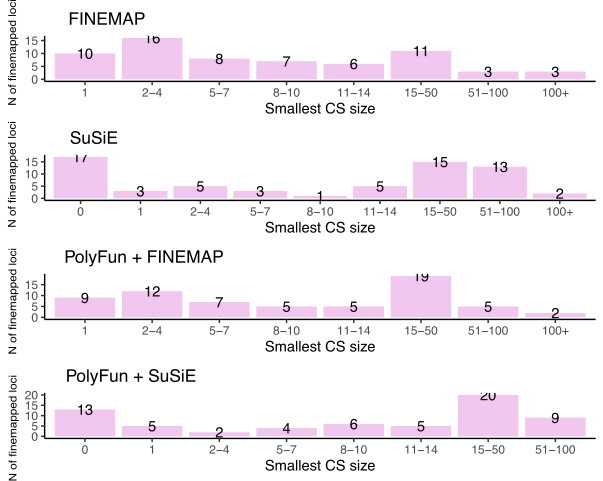

*C). In-sample LD*

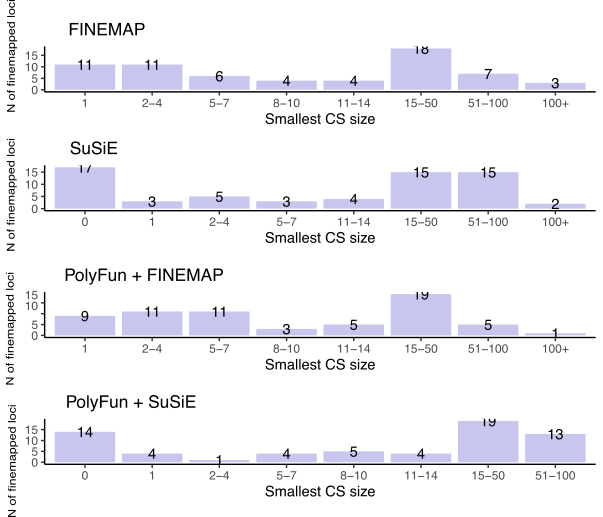

**Figure S4.** **Plot of SNPs with PIP >0.50 across all 16 fine-mapping analyses, including different LD reference panels and fine-mapping methods.** The color of the points corresponds to the LD information used: UK Biobank (pink), Haplotype Reference Consortium (blue), in-sample LD (purple) and no LD (single variant fine-mapping) (grey). Small shapes denote SNPs with PIP between 0.50 and 0.95, while the large shapes denote SNPs with PIP above 0.95. Circles indicate statistical fine-mapping methods and squares indicate functional fine-mapping methods. On the y axis, the genome-wide significant locus names as defined in the original GWAS paper are in parenthesis after each SNP.

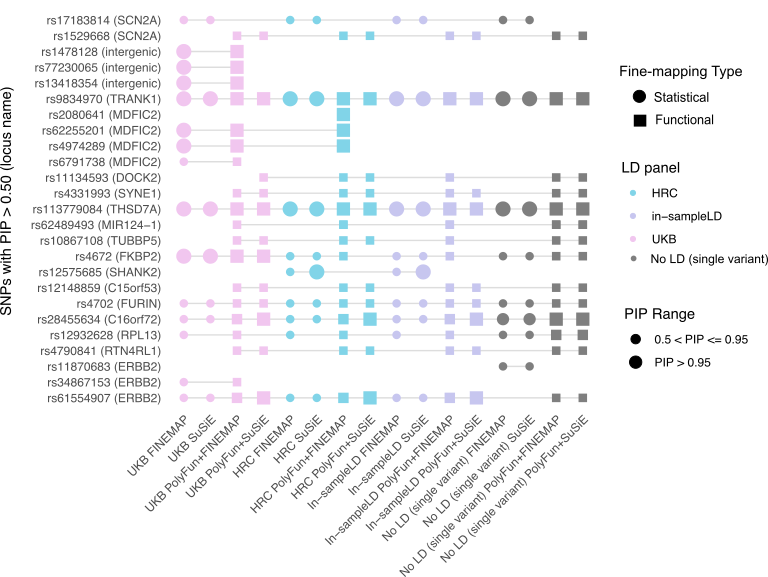

**Figure S5 A-N.** **Multi-track locus plots for fine-mapped loci according to convergence of evidence across validation analyses.** Panels A-N are named according to the genome-wide significant (GWS) locus name assigned in the BD GWAS (Mullins et al., 2021). The upper track depicts the GWAS association statistics over a window of 100,000 base pairs. Variants are colored according to their linkage disequilibrium (LD) r^2^ with the index SNP, calculated based on the HRC reference panel. In the following four tracks, posterior inclusion probabilities (PIPs) are provided from SuSiE, PolyFun+SuSiE, FINEMAP and PolyFun+FINEMAP based on the Haplotype Reference Consortium LD reference panel. Each bar represents a variant in a credible set, and the bars are colored according to the credible set they belong to. The lower tracks visualize overlap with neuronal enhancers or promoters (Nott et al., 2019) and the gene tracks. On each track, the SNPs labeled represent the SNP prioritized through fine-mapping and the index SNP in the GWS locus. All genomic coordinates are in GRCh37.

A). **TUBBP5 locus** (Chromosome 9) (SNP with PIP> 0.50 and part of a 95% CS, SMR eQTL, sQTL and mQTL evidence)

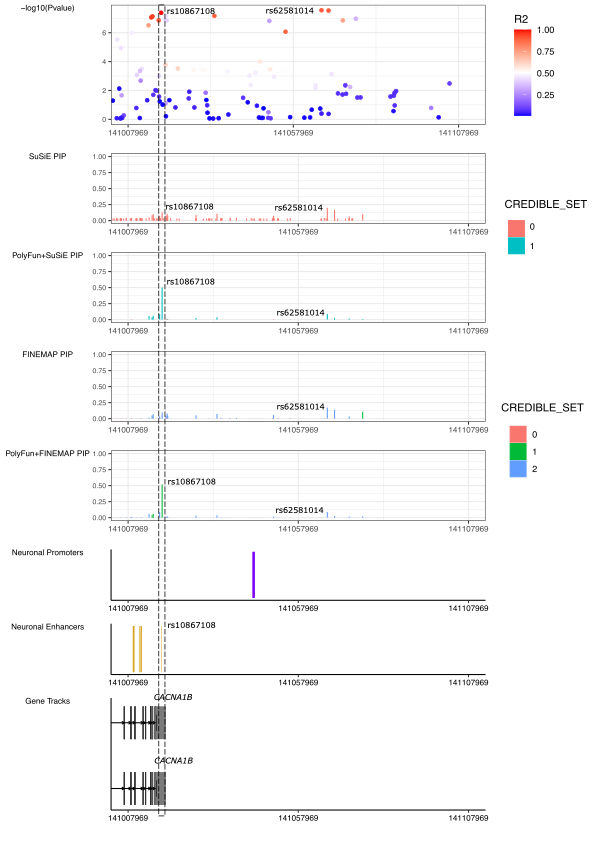

B). **FKBP2 locus** (Chromosome 11) (SNP with PIP> 0.50 and part of a 95%CS, missense variant, SMR eQTL, sQTL and mQTL evidence, overlaps within astrocyte and neuronal promoters)

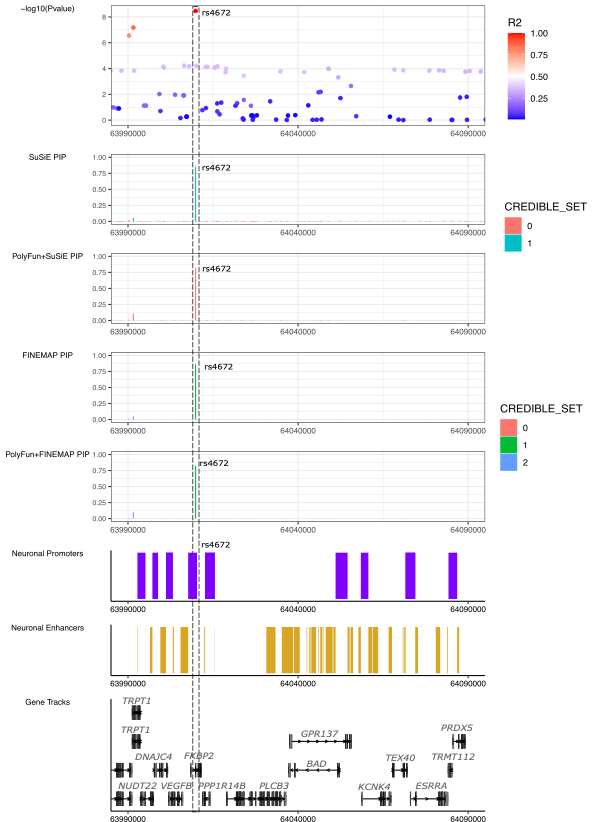

C). **FURIN locus** (Chromosome 15) (SNP with PIP> 0.50 and part of a 95%CS, SMR eQTL, sQTL and mQTL evidence, enhancer-promoter interaction through PLAC-seq)

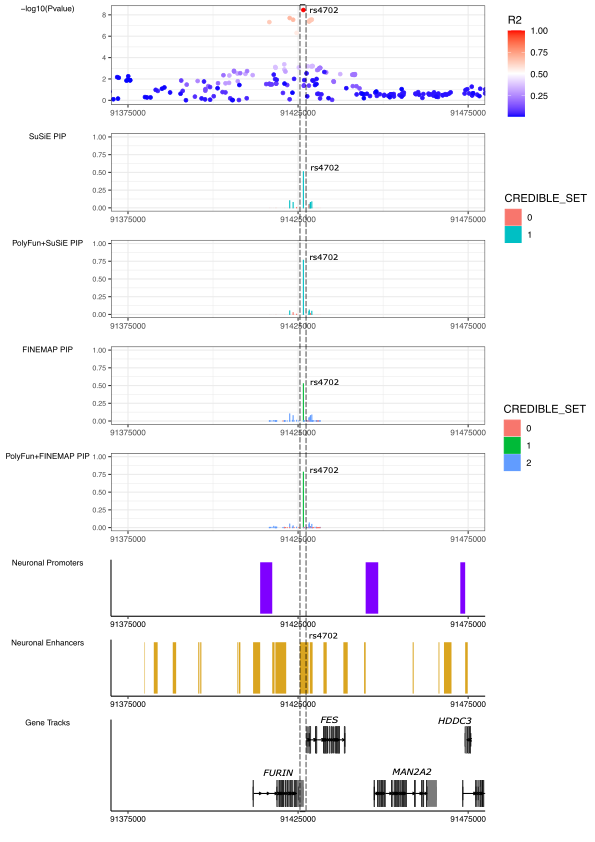

D). **TRANK1 locus** (Chromosome 3) (SNP with PIP> 0.50 and part of a 95%CS, SMR eQTL evidence, overlap within neuronal enhancer)

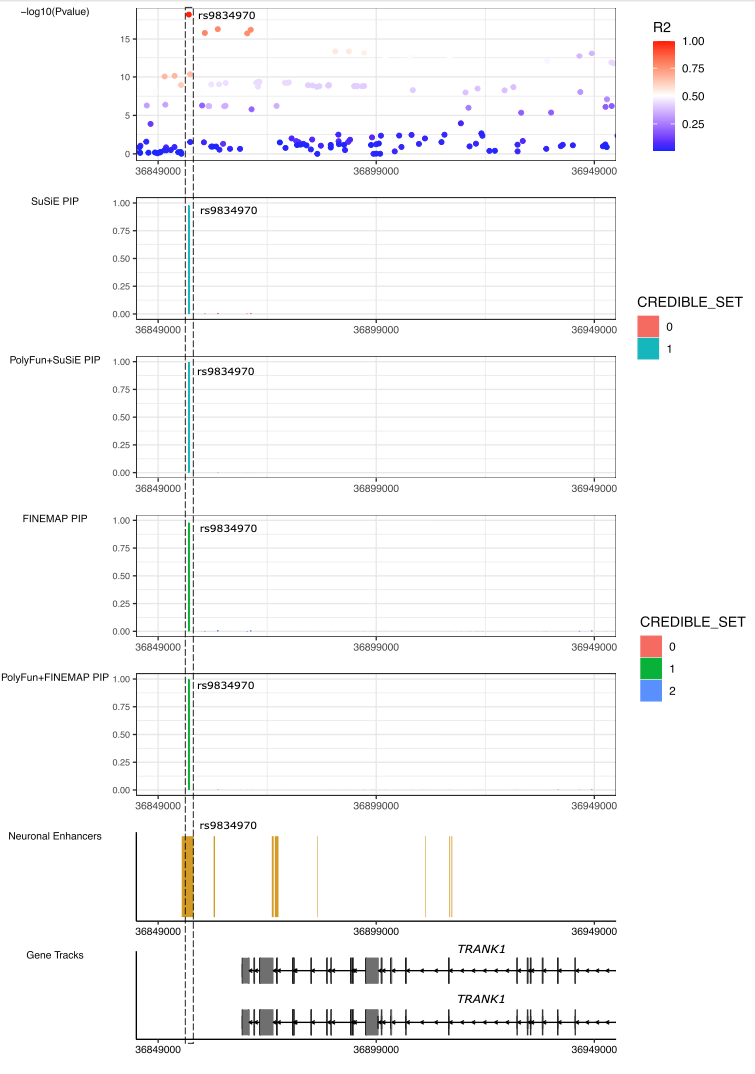

E). **SCN2A locus** (Chromosome 2)(SNP with PIP> 0.50 and part of a 95%CS, missense variant, SMR sQTL evidence, overlap within neuronal promoter)

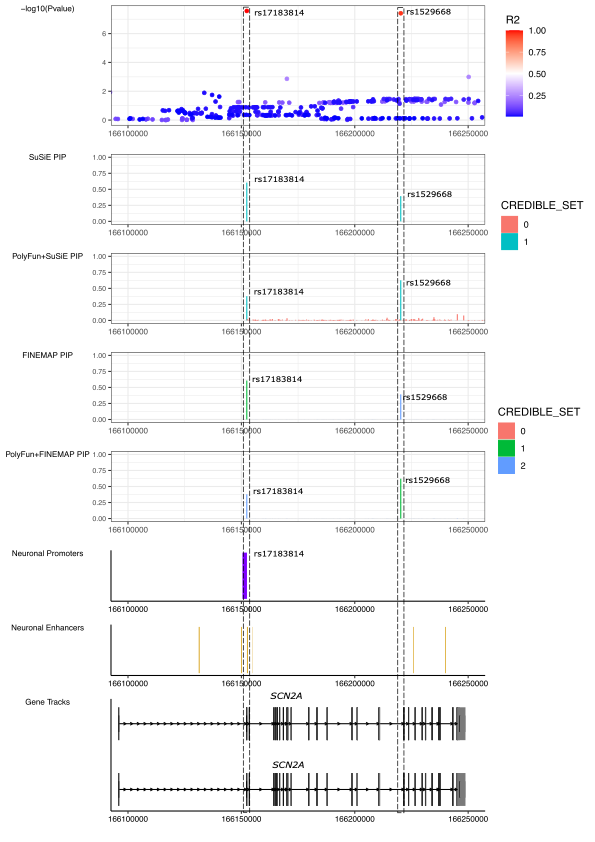

F). **SYNE1 locus** (Chromosome 6) (SNP with PIP> 0.50 and part of a 95%CS, enhancer-promoter interaction through PLAC-seq)

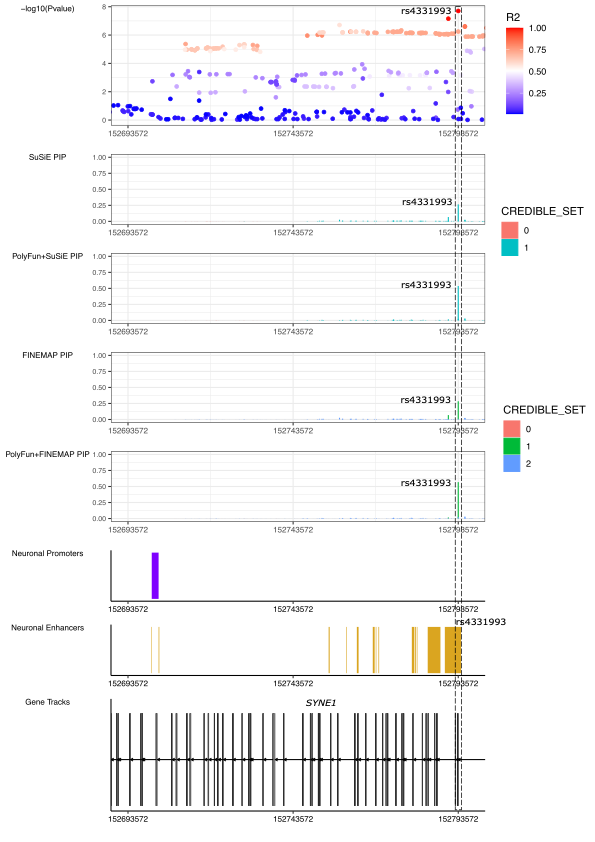

G). **THSD7A locus** (Chromosome 7) (SNP with PIP> 0.50 and part of a 95%CS, overlap within astrocyte and neuronal promoters)

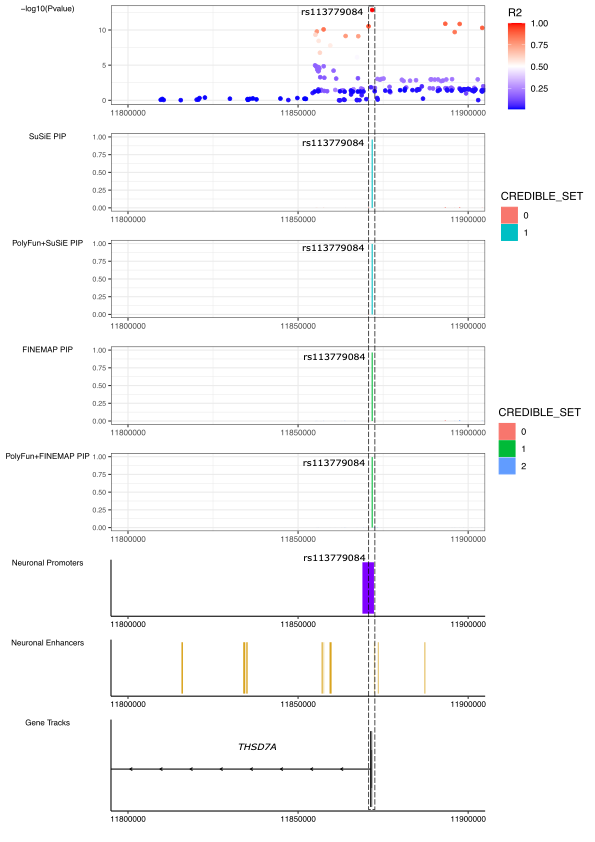

H). **SHANK2 locus** (Chromosome 11; rs12575685) (SNP with PIP> 0.50 and part of a 95%CS)

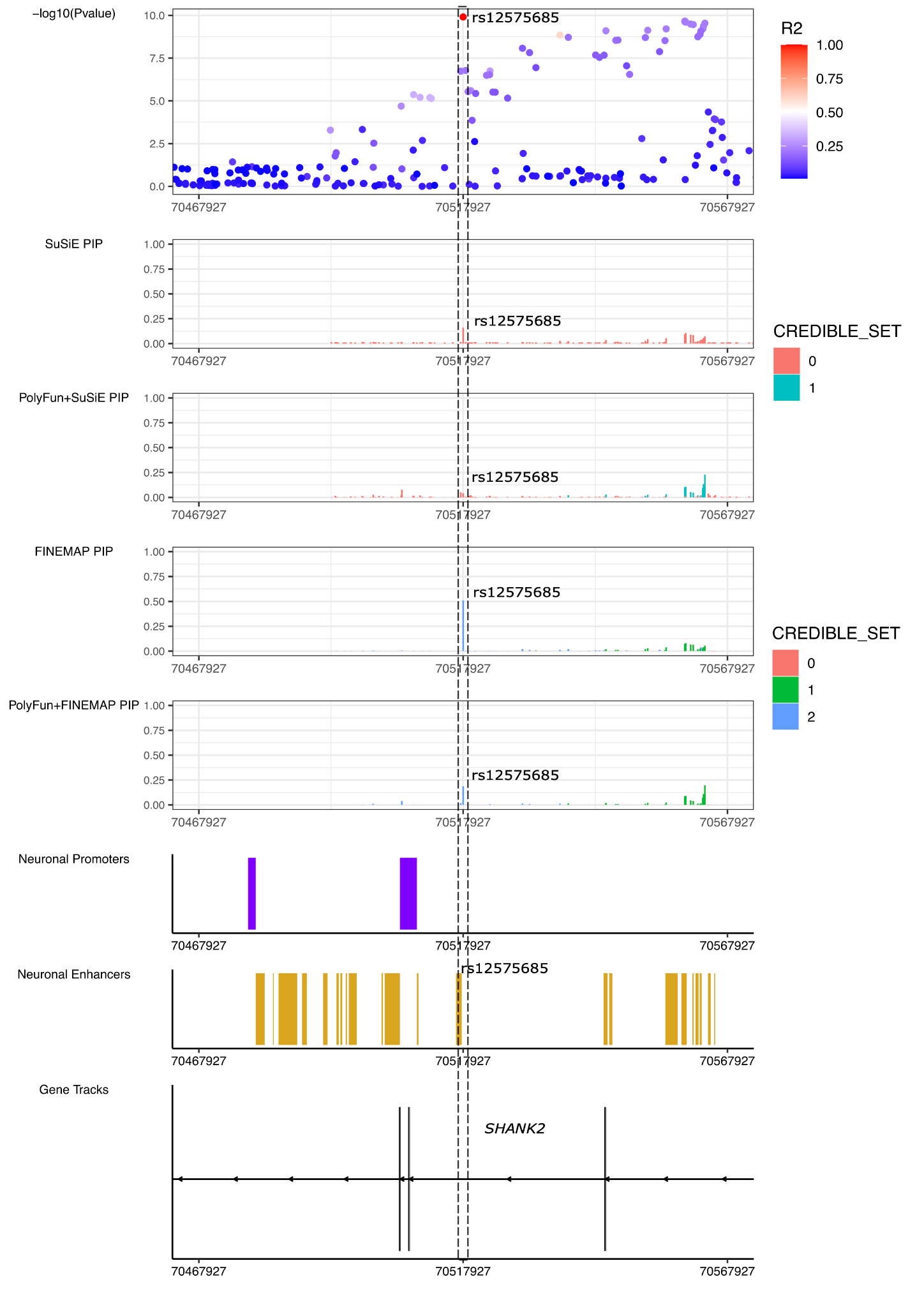

I).**ERBB2 locus** (Chromosome 17; rs61554907) (SNP with PIP> 0.50 and part of a 95%CS, overlap within astrocyte and neuronal promoters)

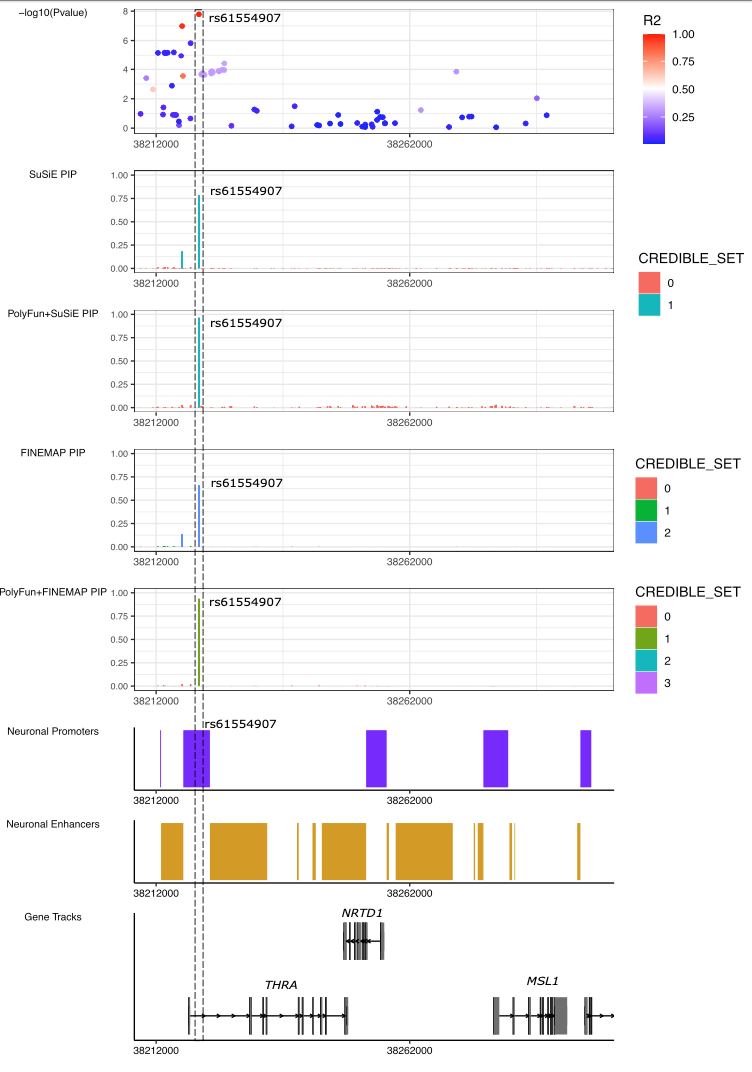

J).**ERBB2 locus** (Chromosome 17; rs11870683) (SNP with PIP> 0.50 and part of a 95%CS, SMR sQTL and mQTL evidence)

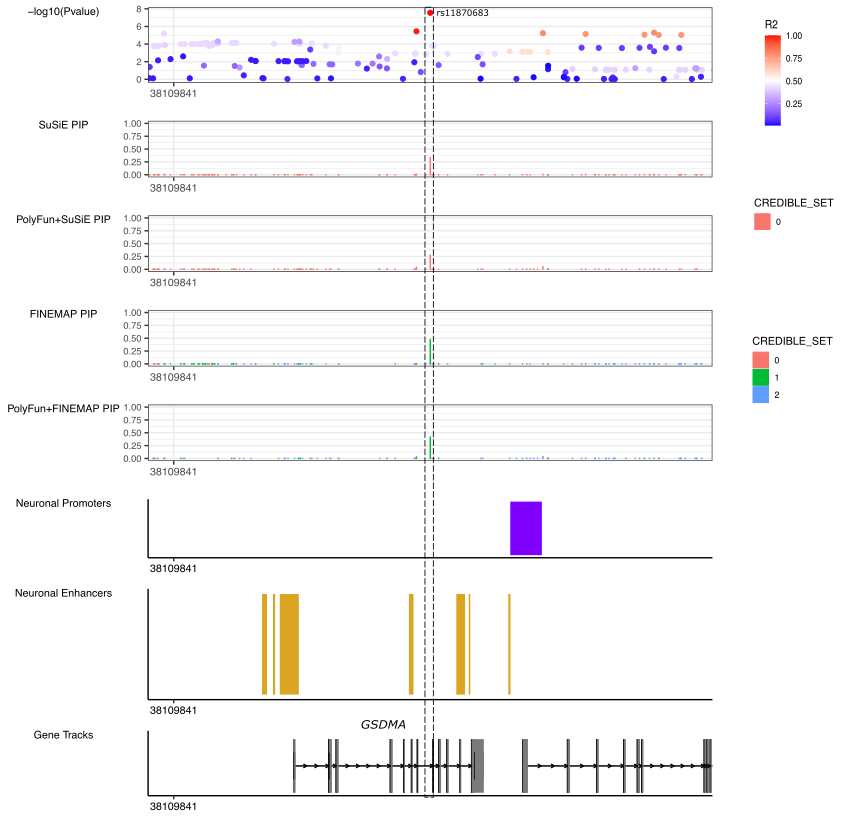

K).**C15orf53 locus** (Chromosome 15) (SNP with PIP> 0.50 and part of a 95%CS, enhancer-promoter interaction through PLAC-seq, overlap within neuronal enhancers)

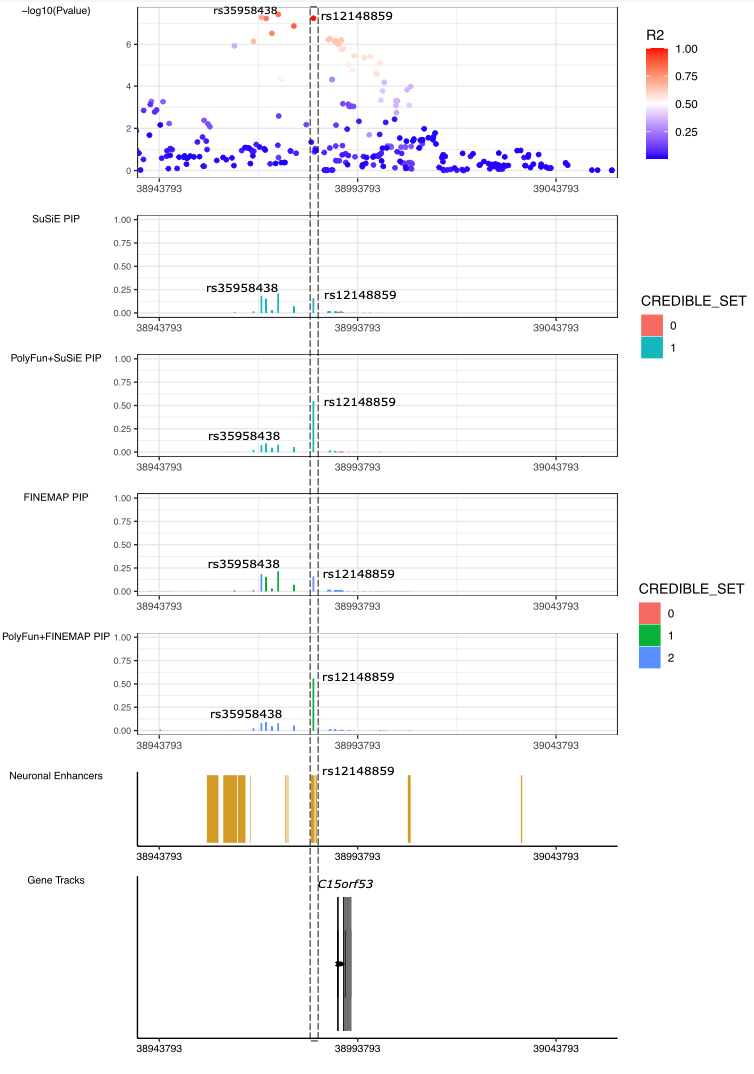

L).**RPL13 locus** (Chromosome 16) (SNP with PIP> 0.50 and part of a 95%CS)

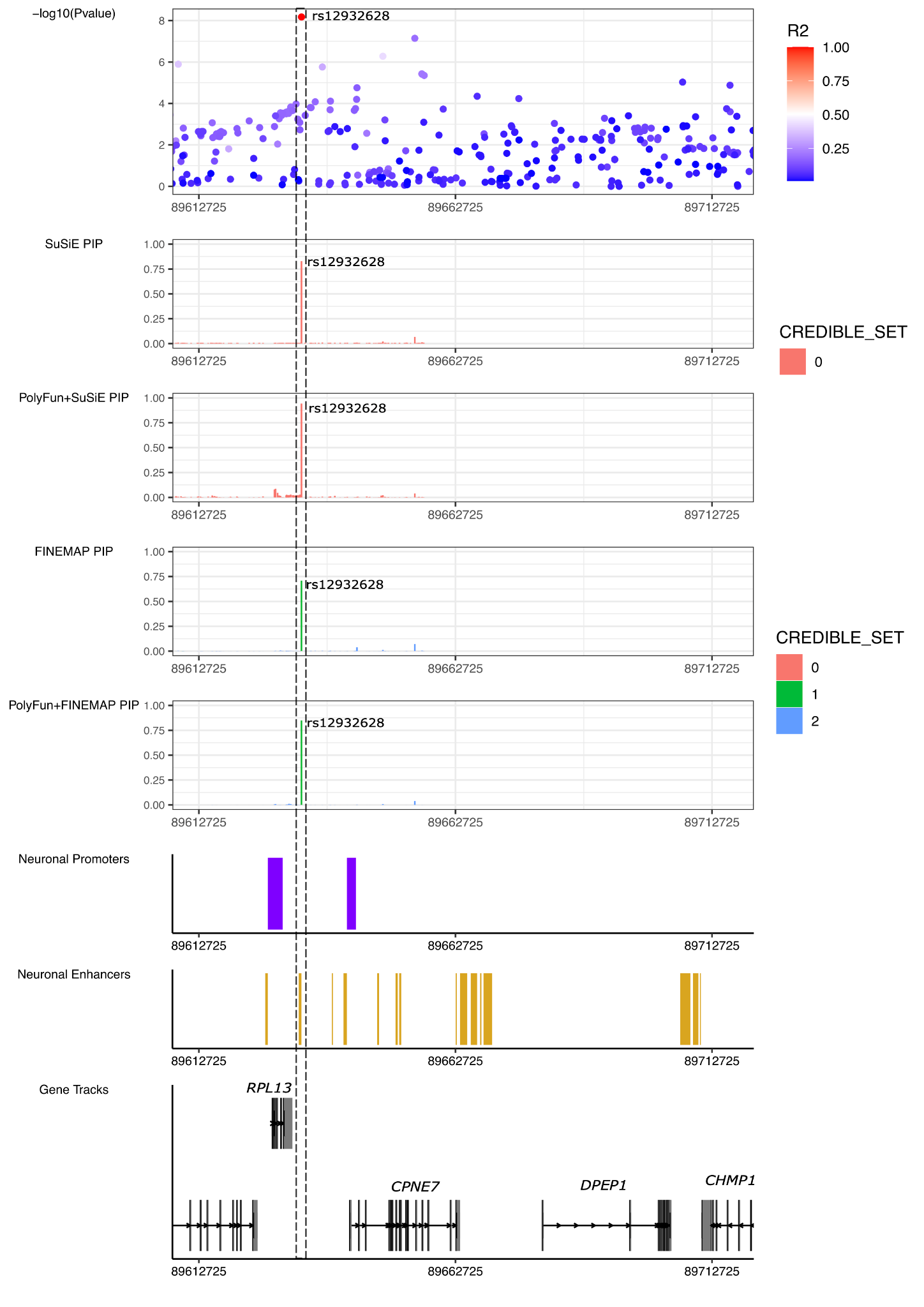

M)**. DOCK2 locus (Chromosome 5)** (SNP with PIP> 0.50 and part of a 95%CS, enhancer-promoter interaction through PLAC-seq, overlap within neuronal enhancers)

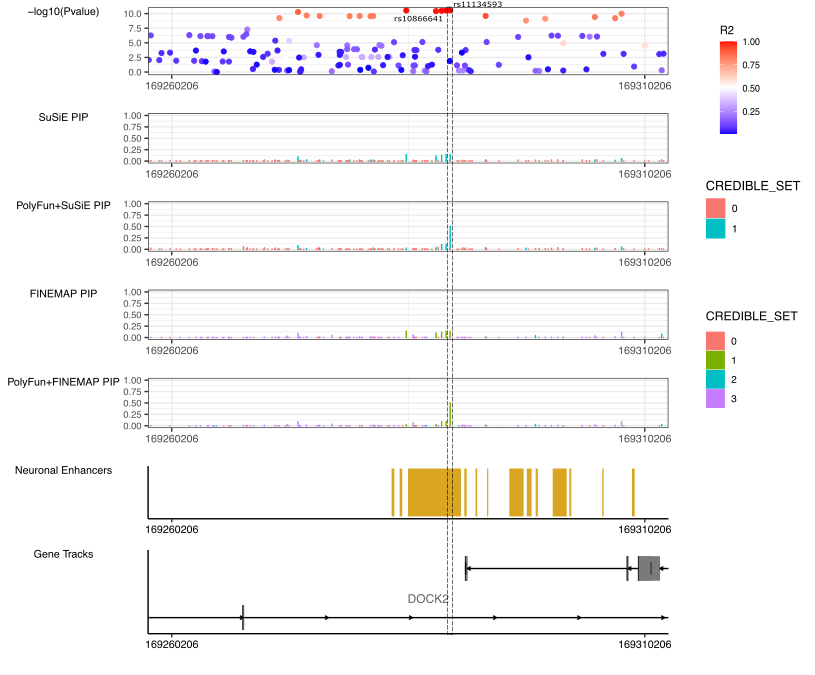

N)**. C16orf72 locus (Chromosome 16)** (SNP with PIP> 0.50 and part of a 95%CS)

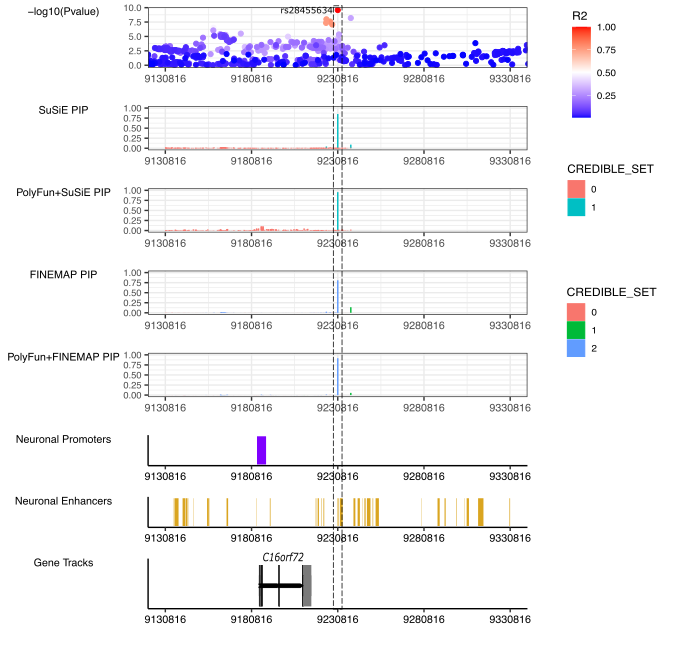

**Figure S6. Area plots of the MHC locus after imputation of HLA variants and amino acids based on the 1000 genomes Phase 3 (European ancestry) reference panel.** Panel A shows the association results from the meta-analysis of 48 BD cohorts before conditioning on the top lead SNP, while panel B shows the association results after conditioning on the top lead SNP (rs1541269). The color of the variants corresponds to their linkage disequilibrium r^2^ value with the index variant in each panel. The shape of each point corresponds to the type of variant - SNP (circle), amino acid (square) or HLA allele (diamond).

*A).*

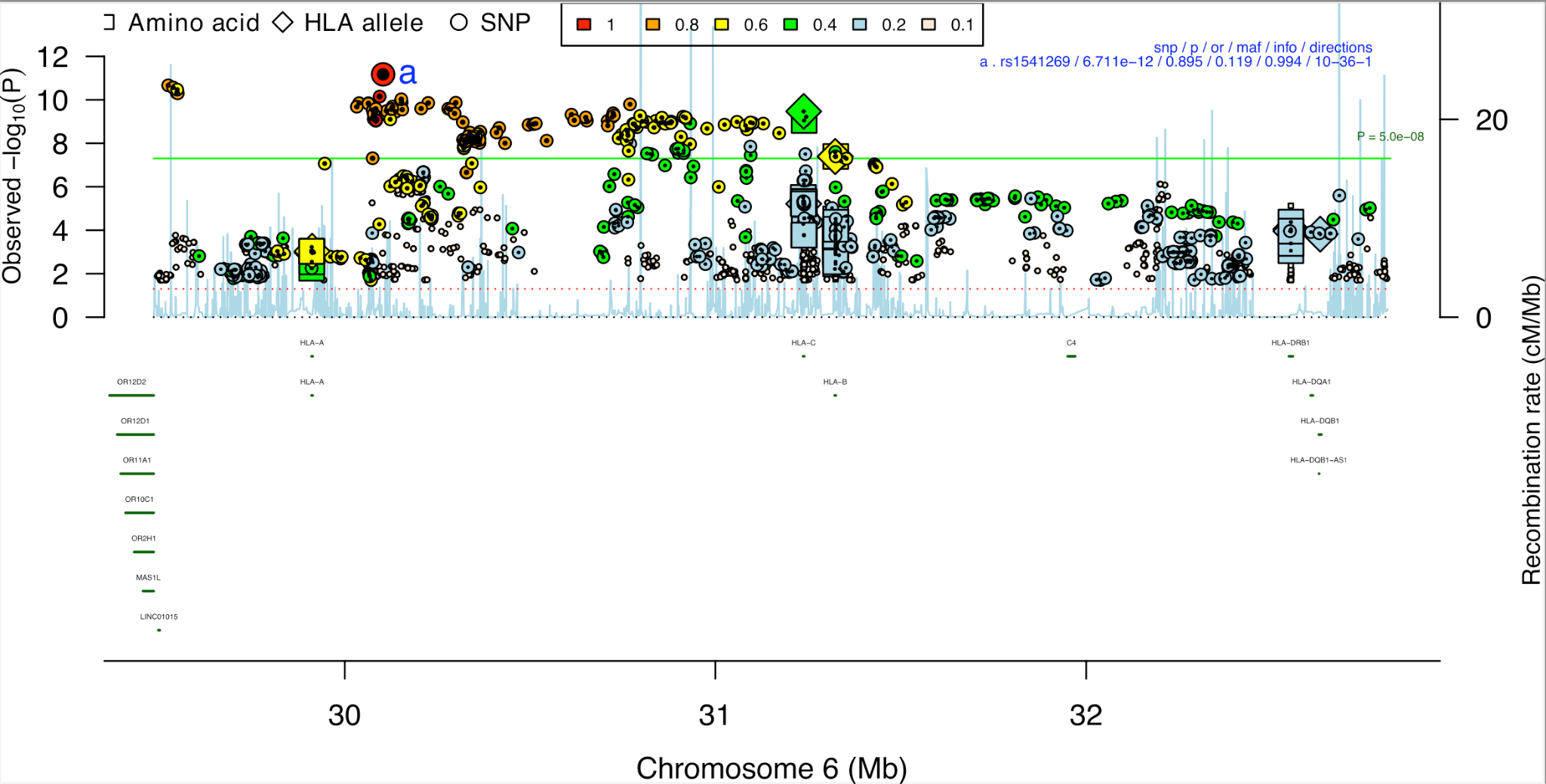

*B).*

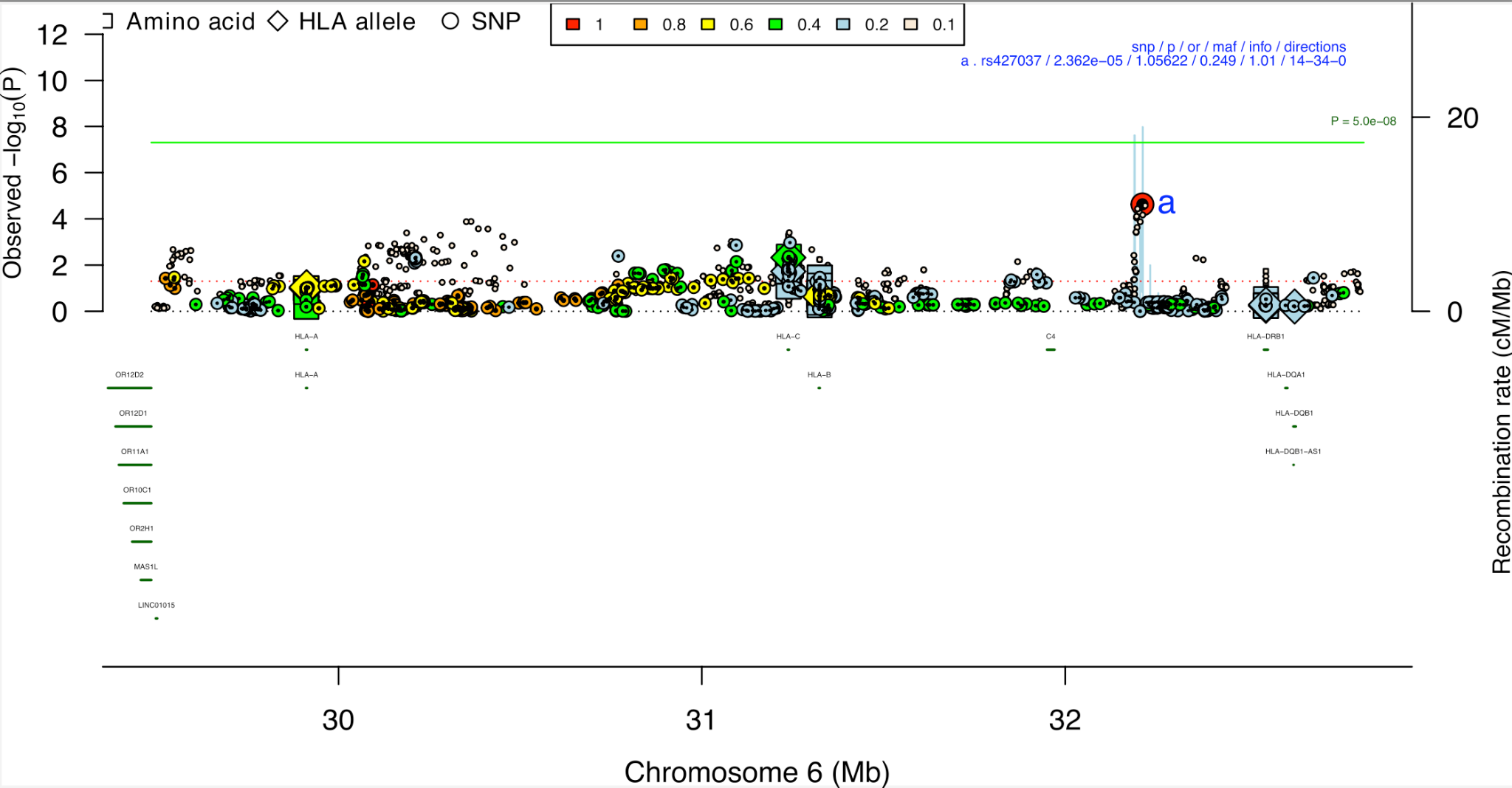

### **Supplementary Note**

#### **Comparison of the impact of LD reference panels on fine-mapping**

We aimed to investigate the impact of using an LD reference panel during fine-mapping, compared with using LD information calculated from the original GWAS data. The latter is typically considered the gold-standard approach, however is difficult in practice due to data availability and sharing restrictions.

We had access to the individual-level genetic data for 48/57 cohorts in the PGC BD GWAS, comprising 33,781 BD cases and 53,869 controls. First, we performed a meta-analysis of the GWAS summary statistics for these 48 cohorts (“meta-48”) using the same procedures as the original GWAS^1^, followed by cleaning of the GWAS results using DENTIST software. These “meta-48” results comprised 7,616,190 SNPs, and there were 45 GWS loci. We calculated “in-sample” LD using the individual-level genetic data from these 48 cohorts, as described in the main text Methods section on “LD reference panels”. We fine-mapped 44 GWS loci (excluding the MHC locus), using the GWAS summary statistics from the “meta-48”, using 4 fine-mapping methods, the GWS locus window and a 3Mb window, using “in-sample” LD calculated based on the exact same set of individuals in the “meta-48” GWAS. These analyses were repeated with the three other LD options: 1) the HRC reference panel, 2) the UKB reference panel and 3) single variant fine-mapping with no LD. The results of each of these fine-mapping analyses were compared against the corresponding analysis based on the “in-sample” LD, by calculating Jaccard Indices of fine-mapped SNPs (again defined as SNPs with PIP > 0.5 and in 95% credible sets). We computed the Jaccard Index across the different LD panels and windows and compared them against those derived from in-sample LD. We observed high levels of concordance between the fine-mapping results based on in-sample LD compared to the HRC (mean Jaccard Index 0.94 (sd 0.12)), followed by UKB (mean Jaccard Index 0.84 (sd 0.10)), and a lesser degree of concordance with the no LD (single variant) fine-mapping (mean Jaccard Index 0.67 (sd 0.05)) (**Supplementary Note Table 1**).

| **Supplementary Note Table 1: Jaccard indices comparing fine-mapping results based on different LD options with those based on “in-sample” LD.** Fine-mapping was performed on the “meta48 GWAS”. Analyses include 4 fine-mapping methods, 2 fine-mapping windows and 3 LD options compared against in-sample LD. Results are based on fine-mapped SNPs with PIP > 0.50 and parts of 95% credible sets. Brackets after each Jaccard index denote the number of SNPs featuring in both sets (intersection), divided by the unique number of SNPs across both sets (union). | | | |
| --- | --- | --- | --- |
| **Fine-mapping window_method** | **HRC** | **UKB** | **No LD (single variant)** |
| GWS locus FINEMAP | 1 (7/7) | 0.88 (7/8) | 0.71 (5/7) |
| GWS locus SuSiE | 1 (7/7) | 1 (7/7) | 0.71 (5/7) |
| GWS locus PolyFun+FINEMAP | 1 (8/8) | 0.88 (8/9) | 0.67 (6/9) |
| GWS locus PolyFun+SuSiE | 1 (6/6) | 0.86 (6/7) | 0.63 (5/8) |
| 3Mb FINEMAP | 0.71 (5/7) | 0.66 (4/6) | 0.6 (3/5) |
| 3Mb SuSiE | 1 (4/4) | 0.8 (4/5) | 0.75 (3/4) |
| 3Mb PolyFun+FINEMAP | 0.78 (7/9) | 0.875 (7/8) | 0.625 (5/8) |
| 3Mb PolyFun+SuSiE | 1 (4/4) | 0.75 (3/4) | 0.67 (4/6) |
| **Mean (sd)** | **0.94 (0.12)** | **0.84 (0.10)** | **0.67 (0.05)** |

Second, we calculated the correlation of the PIP values of SNPs fine-mapped (PIP > 0.5 and parts of 95% credible sets) using both the in-sample LD and the specified LD option (HRC, UKB and no LD (single variant fine-mapping) (**Supplemental Note Table 2**). Amongst SNPs fine-mapped by both in-sample LD and each specified LD option, PIP values were extremely highly correlated.

| **Supplementary Note Table 2: Correlation (r) of posterior inclusion probability (PIP) values of SNPs fine-mapped using both the in-sample LD and the specified LD option.** Fine-mapping was performed on the “meta48 GWAS”. Analyses include 4 fine-mapping methods, 2 fine-mapping windows and 3 LD options compared against in-sample LD. Results are based on fine-mapped SNPs with PIP > 0.50 and parts of 95% credible sets. Values in brackets after each correlation denote the number of SNPs fine-mapped using both the specified LD option and in-sample LD. | | | |
| --- | --- | --- | --- |
| **Fine-mapping window_method** | **HRC** | **UKB** | **No LD (single variant)** |
| GWS locus FINEMAP | 0.998 (7) | 0.999 (7) | 0.970 (5) |
| GWS locus SuSiE | 0.999 (7) | 0.999 (7) | 0.987 (5) |
| GWS locus PolyFun+FINEMAP | 0.994 (8) | 0.989 (8) | 0.998 (6) |
| GWS locus PolyFun+SuSiE | 0.999 (6) | 0.999 (6) | 0.999 (5) |
| 3Mb FINEMAP | 0.999 (5) | 0.999 (4) | 0.999 (3) |
| 3Mb SuSiE | 0.999 (4) | 0.997 (4) | 0.999 (3) |
| 3Mb PolyFun+FINEMAP | 0.999 (7) | 0.999 (7) | 0.920 (5) |
| 3Mb PolyFun+SuSiE | 0.999 (4) | 0.997 (3) | 0.902 (4) |
| **Mean (sd)** | **0.998 (0.002)** | **0.997 (0.003)** | **0.972 (0.039)** |

Third, we assessed the correlation of the allele frequencies calculated from the “in-sample” LD dataset and the HRC reference panel. Between the HRC and in-sample LD (7,611,652 SNPs from the “meta48” GWAS summary statistics found in both), the correlation of allele frequencies was 0.99, further emphasizing the comparability of the two datasets. Allele frequency calculations were not possible for the UK Biobank LD reference panel used, as data were provided as LD matrices rather than individual-level genetic data.

In summary, analyses of the impact of LD reference panels on fine-mapping demonstrate the strong concordance of results between the HRC reference panel and the in-sample LD, supporting our hypothesis that a well-matched LD reference panel to the GWAS summary statistics can achieve high fine-mapping resolution, when in-sample LD estimates are not available.

#### **Comparison of the impact of locus windows on fine-mapping**

To assess the impact of locus windows on fine-mapping, we fine-mapped the 44 GWS loci from the meta48 GWAS using the GWS locus window (defined as in the original GWAS as LD r^2^ > 0.1 with the top lead SNP within a 3 Mb range) and a 3Mb fine-mapping window. The 3 Mb fine-mapping windows were selected using the 2,763 published LD matrices of 3 Mb blocks covering the entire genome calculated in the UKB, such that the index SNP was as close as possible to the center of the 3Mb window. We used the coordinates of these 3 Mb windows, as they are widely used to obtain LD information for post-GWAS analyses, are provided for download with the Polyfun and PolyPred softwares, and represent a large (n=337,000) publicly available European ancestry LD reference panel. Keeping the LD option and fine-mapping method consistent, we calculated the Jaccard Indices of fine-mapped SNPs (defined as SNPs with PIP > 0.5 and in 95% credible sets) between the GWS locus and 3Mb windows (**Supplemental Note Table 3).** Comparing the two fine-mapping windows, Jaccard Indices ranged from 0.43 to 0.86 across analyses, with mean 0.59 (sd 0.12).

| **Supplementary Note Table 3: Jaccard indices comparing fine-mapping results from GWS locus versus 3Mb fine-mapping windows.** Fine-mapping was performed on the “meta48 GWAS”. Analyses include 4 fine-mapping methods and 4 LD options and 2 fine-mapping windows were compared. Results are based on fine-mapped SNPs with PIP > 0.50 and parts of 95% credible sets. Brackets after each Jaccard Index denote the number of SNPs featuring in both sets (intersection), divided by the unique number of SNPs across both sets (union). | |
| --- | --- |
| **LD option_Fine-mapping method** | **GWS locus vs. 3Mb fine-mapping window** |
| In-sample LD FINEMAP | 0.5 (4/8) |
| In-sample LD SuSiE | 0.57 (4/7) |
| In-sample LD PolyFun+FINEMAP | 0.5 (5/10) |
| In-sample LD Polyfun+SuSiE | 0.5 (4/8) |
| HRC FINEMAP | 0.55 (5/9) |
| HRC SuSiE | 0.57 (4/7) |
| HRC PolyFun+FINEMAP | 0.54 (6/11) |
| HRC Polyfun+SuSiE | 0.5 (4/8) |
| UKB FINEMAP | 0.625 (5/8) |
| UKB SuSiE | 0.714 (5/7) |
| UKB PolyFun+FINEMAP | 0.55 (6/11) |
| UKB Polyfun+SuSiE | 0.43 (3/7) |
| No LD (single variant) FINEMAP | 0.6 (3/5) |
| No LD (single variant) SuSiE | 0.6 (3/5) |
| No LD (single variant) PolyFun+FINEMAP | 0.857 (6/7) |
| No LD (single variant) Polyfun+SuSiE | 0.857 (6/7) |
| **Mean (sd)** | **0.59 (0.12)** |

We calculated the correlation of the PIP values of SNPs fine-mapped (PIP > 0.5 and parts of 95% credible sets) using both the GWS locus and 3 Mb fine-mapping window (**Supplemental Note Table 4**). Despite the modest Jaccard Indices of concordance in fine-mapping results between the two fine-mapping windows, the PIP correlation values between SNPs successfully fine-mapped using both fine-mapping windows were high (mean PIP correlation 0.988 (sd 0.025)).

| **Supplementary Note Table 4: Correlation (r) of posterior inclusion probability (PIP) values of fine-mapped SNPs identified in both fine-mapping windows (GWS locus vs 3Mb)**. These results are based on GWAS meta-analysis using only the 48 cohorts for which we have individual level genetic data available. The analyses include 4 LD options and 4 fine-mapping methods, and they are based on fine-mapped SNPs with PIP > 0.50 and parts of 95% credible sets. Values in the brackets denote the number of shared fine-mapped SNPs amongst the different LD panels and in-sample LD. | |
| --- | --- |
| **LD option_Fine-mapping method** | **GWS locus vs. 3Mb fine-mapping window** |
| In-sample LD FINEMAP | 0.997 (4) |
| In-sample LD SuSiE | 0.997 (4) |
| In-sample LD PolyFun+FINEMAP | 0.999 (5) |
| In-sample LD Polyfun+SuSiE | 0.999 (4) |
| HRC FINEMAP | 0.997 (5) |
| HRC SuSiE | 0.998 (4) |
| HRC PolyFun+FINEMAP | 0.995 (6) |
| HRC Polyfun+SuSiE | 0.999 (4) |
| UKB FINEMAP | 0.997 (5) |
| UKB SuSiE | 0.997 (5) |
| UKB PolyFun+FINEMAP | 0.995 (6) |
| UKB Polyfun+SuSiE | 0.997 (3) |
| No LD (single variant) FINEMAP | 0.999 (3) |
| No LD (single variant) SuSiE | 0.999 (3) |
| No LD (single variant) PolyFun+FINEMAP | 0.926 (6) |
| No LD (single variant) Polyfun+SuSiE | 0.921 (6) |
| **Mean (sd)** | **0.988 (0.025)** |

In summary, our comparison of the impact of LD reference panels and fine-mapping windows on the “meta48” GWAS, indicated that fine-mapping results were more similar when using different LD options (**Supplementary Note Tables 1 and 2**) than a GWS locus versus a 3Mb fine-mapping window (**Supplementary Note Tables 3 and 4**). For the full PGC3 BD GWAS, we ran fine-mapping using the four LD options to ensure the robustness of fine-mapped SNPs. In the full PGC3 BD GWAS, 3 of the GWS loci were larger than 3Mb, so could not be fully covered by a 3Mb window. The remaining 61 GWS loci varied greatly in size, ranging from 14,960 bp - 1.96 Mb (mean 375 kb (sd 390 kb). Based on these results, we fine-mapped the GWS loci from the full PGC3 BD GWAS using only the GWS locus fine-mapping windows (defined in the original GWAS as LD r^2^ > 0.1 with the top lead SNP within a 3 Mb range), as these best reflect the regions of genome-wide significant association from the GWAS. Additionally, using a fine-mapping window that is significantly larger than the GWS locus window, may fine-map other nearby regions that capture different association signals to that represented by the GWS index SNP.

#### **Description of testing cohorts used for polygenic risk scoring analyses**

Below we describe the ascertainment and diagnosis of the participants in each testing cohort. Most cohorts have been published individually, and the primary report can usually be found using the PubMed identifiers provided. The lead PI of each sample warranted that their protocol was approved by their local Ethical Committee and that all participants provided written informed consent. The boldfaced first line for each sample indicates study PI, PubMed ID if published, country (study name), the Psychiatric Genomics Consortium internal tag or study identifier, and genetic ancestry.

**Grigoroiu-Serbanescu M | PMID : 31791676| Romania (BOMA-Romania) | rom4 | European**

**Patient sample.** Unrelated bipolar disorder type I (BD-I) patients (N=102) were recruited from consecutive admissions in the Obregia Psychiatric Hospital of Bucharest, Romania. All participants provided written informed consent following a detailed explanation of the study aims and procedures. The study was performed in accordance with the Code of Ethics of the World Medical Association (Declaration of Helsinki) and it was approved by the ethical committee of the hospital and by the reviewers of the Research and Education Ministry. All participants were of Romanian descent according to self-reported ancestry. Genealogical information about parents and all four grandparents was obtained through direct interview of the subjects. The patients were investigated with the Diagnostic Interview for Genetic Studies (DIGS)^2^ and the Family Interview for Genetic Studies (FIGS)^3^. Information was also obtained from medical records and close relatives. The diagnosis of BD-I was assigned according to DSM-IV-TR criteria using the best estimate procedure that considered patient interview, medical records and information provided by close relatives. Patients were included in the sample if they had at least two documented hospitalized illness episodes (one manic/mixed and one depressive or two manic episodes) and no residual mood incongruent psychotic symptoms during remissions. This information was also confirmed by first degree relatives for 78% of the cases in face to face interviews. Family history of psychiatric illness was obtained with FIGS administered both to the patients and to all available first degree relatives.

**Control sample**. The controls (N=198) were volunteers from the personnel and students of the University of Bucharest, as well as from personnel and medical residents at the Obregia Psychiatric Hospital and the Institute of Virology of Bucharest. All controls were evaluated using the DIGS and FIGS to screen for a lifetime history of major affective disorders, schizoaffective disorders, SCZ and other psychoses, obsessive-compulsive disorder, eating disorders, and alcohol or drug addiction. Unaffected individuals were included as controls in the present study.

**Genotyping of the Romanian patients and controls**. The BD-I cases and controls were genome-wide genotyped on Illumina GSA-MD beadchips. Stringent quality control was applied to the genotype information. Individuals were excluded on the basis of having incorrect gender assignments; excessive heterozygosity (more than 10 standard deviations above the mean); missing genotype data above 10% and evidence of relatedness. SNPs were excluded with a minor allele frequency < 0.5% and deviating substantially from the Hardy-Weinberg equilibrium (P < 10^−6^).

**Kircher T | PMID 30267149| Germany | FOR 2107 |European**

The FOR2107 cohort is a multi-centre study, recruited through newspaper advertisements and mailing lists from the areas of Marburg and Muenster in Germany^4^. The sample includes 147 cases and 696 controls, genotyped with the PsychChip platform. Ethics approval was obtained from the ethics committees of the Medical Schools of the Universities of Marburg and Muenster, respectively, in accordance with the Declaration of Helsinki. All subjects volunteered to participate in the study and provided written informed consent.

**McQuillin A | PMID: 37643680 | UCL (University College London), London, UK | amq1| European**

The Amq1 cohort is a study taking place in London, United Kingdom. Diagnostic criteria were based on ICD-10 codes and clinical interviews and controls were screened for any mental disorder. The cohort includes 417 cases (201 bipolar disorder type I (BD-I) and 39 bipolar disorder type II (BD-II) cases) and 533 control samples, genotyped using the A5.0 platform. The sample was composed of Caucasian individuals who were ascertained and received clinical diagnoses of BD-I according to UK National Health Service (NHS) psychiatrists at interview using ICD-10 codes. In addition bipolar subjects were included only if both parents were of English, Irish, Welsh or Scottish descent and if three out of four grandparents were of the same descent. All volunteers read an information sheet approved by the Metropolitan Medical Research Ethics Committee who also approved the project for all NHS hospitals. Written informed consent was obtained from each volunteer. The control subjects were recruited from London branches of the National Blood Service, from local NHS family doctor clinics and from university student volunteers. All control subjects were interviewed with the SADS-L to exclude all psychiatric disorders.

**Pato M, Pato C, Bigdeli T | PMID: 33169155 | USA | GPC | Admixed African American, Latino**

Details of ascertainment and diagnosis, genotyping and quality control have been described in detail previously^5^. Briefly, cases were ascertained using the Diagnostic Interview for Psychosis and Affective Disorders (DI-PAD), a semi-structured clinical interview administered by mental health professionals, which was developed specifically for the GPC study. Individuals reporting no lifetime symptoms indicative of psychosis or mania and who have no first-degree relatives with these symptoms are included as control participants. Genotyping of the cohort was performed in 7 ‘batches’ using Illumina Infinium arrays (Omni2.5, Multi-Ethnic Global Array, and Global Screening Array). Typed variants were aligned to the human reference genome (GRCh37), and within each genotyping batch, variants with missingness greater than 2% or Hardy-Weinberg Equilibrium *P*-value<10-6 were excluded; all scripts for pre-processing GWAS array data are downloadable from https://github.com/freeseek/gwaspipeline.

Computational phasing and statistical genotype imputation were performed for each genotyping batch using Eagle (v2.3.5) and Minimac3 (v2.0.1), respectively, with default parameters and using publicly available reference haplotypes from the 1000 Genomes Project (1KGP) Phase 3. Principal components analysis (PCA) was performed with GCTA (v1.2.4), using a genome-wide genetic relatedness matrix (GRM) estimated for the full GPC dataset and reference samples from the 1KGP Phase 3 data based on 34,918 genotyped SNPs. For each individual, we estimated genome-wide average proportions of African (AFR), European (EUR), Admixed American (AMR), East Asian (EAS), and South Asian (SAS) ancestry from global ancestry PCs using a simple linear mixed model. The African American GPC cohort included 1766 cases and 2535 controls, while the Latino GPC cohort comprised 1032 cases and 3090 controls.

**Iwata N | PMID: 28115744 | Japan (advanced COSMO and Biobank Japan) | East Asian**

A detailed description of the sample information, genotyping, quality control and imputation procedures is reported elsewhere^6^. In brief, 2,964 BD and 61,887 comparison subjects from the Japanese population were included in this dataset (genotyped by Illumina OmniExpressExome v1.0 or v1.2 BeadChips). After the imputation and stringent QC, a total of 6,195,093 imputed SNPs were analyzed for the association analysis. The diagnosis for each case subject followed the DSM-IV-TR criteria for BD and schizoaffective disorder and was reached by the consensus of at least two experienced psychiatrists, based on unstructured interviews with the subject and their family, as well as a review of the subject's medical records. For the comparison subjects, we used GWAS data for subjects in the BioBank Japan project collected as case subjects for non-psychiatric disorders. These subjects were not psychiatrically evaluated.

**Hong-Hee Won, Woojae Myung, Heon-Jeong Lee, Genoplan Research Team | Not published | South Korea | East Asian**

We genotyped 807 patients with bipolar disorder, 726 patients with schizophrenia and 497 healthy control subjects using the Affymetrix AxiomR Korea Biobank Array 1.0 (K-CHIP). K-CHIP was designed by the Center for Genome Science at the Korea National Institute of Health, including 833K SNPs. A more detailed description of the genotyping procedure is reported elsewhere. We performed sample-level and variant-level QC of genotype data. We excluded variants with missing rate > 1%, Hardy-Weinberg equilibrium P < 10^-6^, or minor allele frequency < 1%, and samples with missing rate > 5%, relatedness among the sample, mismatch between self-reported and inferred sex, or deviated heterozygosity rate. We confirmed homogeneity of the samples based on visual inspection of principal component analysis plots. Genotype imputation was conducted using the Haplotype Reference Consortium (HRC) reference panel. After the imputation and additional post-QC (R^2^ > 0.8 and minor allele frequency > 1%), a total of 770 bipolar cases and 497 controls and 5,483,856 variants were analyzed for polygenic risk score. All the patients met the DSM-IV-TR diagnostic criteria for bipolar I disorder and bipolar II disorder. For clinical diagnosis, a structured interview using the Korean version of the Diagnostic Interview for Genetic Studies (DIGS) or the Structured Clinical Interview for DSM-IV (SCID) was performed. The control group consisted of volunteers from the community who were free of any history of clinically significant psychiatric symptoms. Detailed assessment processes are described elsewhere^7^.

### **Full Acknowledgments**

We thank the participants who donated their time, life experiences and DNA to this research and the clinical and scientific teams that worked with them. This project was funded by the Baszucki Brain Research Fund via the Milken Institute Center for Strategic Philanthropy. We are deeply indebted to the investigators who comprise the PGC. The PGC has received major funding from the US National Institute of Mental Health (PGC4: R01MH124839, PGC3: U01 MH109528; PGC2: U01 MH094421; PGC1: U01 MH085520). Statistical analyses were carried out on the NL Genetic Cluster Computer (<http://www.geneticcluster.org>) hosted by SURFsara and the Mount Sinai high performance computing cluster ([http://hpc.mssm.edu](http://hpc.mssm.edu/)), which is supported by the Office of Research Infrastructure of the National Institutes of Health under award numbers S10OD018522 and S10OD026880. The content is solely the responsibility of the authors and does not necessarily represent the official views of the National Institutes of Health.

**Investigator Acknowledgements**

NM and MK were part funded by the Baszucki Brain Research Fund via the Milken Institute Center for Strategic Philanthropy and the NIMH US National Institute of Mental Health (R01MH124839).

JRIC is part funded by the NIHR Maudsley Biomedical Research Centre. This paper represents independent research funded by the NIHR Maudsley Biomedical Research Centre at South London and Maudsley NHS Foundation Trust and King’s College London. The views expressed are those of the authors and not necessarily those of the UK NIHR or Department of Health and Social Care. JRIC is part funded by a grant from the UK Medical Research Foundation (MRF-001-0012-RG-COLE-C0930).

**Cohort Acknowledgements**

BACCS: This work was supported in part by the NIHR Maudsley Biomedical Research Centre (‘BRC’) hosted at King’s College London and South London and Maudsley NHS Foundation Trust, and funded by the National Institute for Health Research under its Biomedical Research Centres funding initiative. The views expressed are those of the authors and not necessarily those of the BRC, the NHS, the NIHR or the Department of Health or King’s College London. We gratefully acknowledge capital equipment funding from the Maudsley Charity (Grant Reference 980) and Guy’s and St Thomas’s Charity (Grant Reference STR130505).

BACCS-Canada: This work was supported through funding from the Canadian Institutes of Health Research, MOP-172013, to JBV at the Centre for Addiction & Mental Health, Toronto. The ascertainment of the case control cohorts was also supported by funding from GlaxoSmithKline to JBV.

BD_TRS: This work was funded by the German Research Foundation (DFG, grant FOR2107 DA1151/5-1 to UD; SFB-TRR58, Project C09 to UD) and the Interdisciplinary Center for Clinical Research (IZKF) of the medical faculty of Münster (grant Dan3/012/17 to UD).

BiGS, GAIN: FJM was supported by the NIMH Intramural Research Program, NIH, DHHS.

BOMA-Australia: This work was supported by the Australian National Health and Medical Research Council, grant numbers: 1037196 (PBM, PRS), 1066177 (JMF, JIN), 1063960 (JMF, PRS); and the Lansdowne Foundation. BOMA-Germany I, BOMA-Germany II, BOMA-Germany III, PsyCourse: This work was supported by the German Ministry for Education and Research (BMBF) through the Integrated Network IntegraMent (Integrated Understanding of Causes and Mechanisms in Mental Disorders), under the auspices of the e:Med program (grant 01ZX1314A/01ZX1614A to MMN and SC, grant 01ZX1314G/01ZX1614G to MR, grant 01ZX1314K to TGS). This work was supported by the German Ministry for Education and Research (BMBF) grants NGFNplus MooDS (Systematic Investigation of the Molecular Causes of Major Mood Disorders and Schizophrenia; grant 01GS08144 to MMN and SC, grant 01GS08147 to MR). This work was also supported by the Deutsche Forschungsgemeinschaft (DFG), grant NO246/10-1 to MMN (FOR 2107), grant RI 908/11-1 to MR (FOR 2107), grant WI 3429/3-1 to SHW, grants SCHU 1603/4-1, SCHU 1603/5-1 (KFO 241) and SCHU 1603/7-1 (PsyCourse) to TGS. This work was supported by the Swiss National Science Foundation (SNSF, grant 156791 to SC). MMN is supported through the Excellence Cluster ImmunoSensation. TGS is supported by an unrestricted grant from the Dr. Lisa-Oehler Foundation. AJF received support from the BONFOR Programme of the University of Bonn, Germany. MH was supported by the Deutsche Forschungsgemeinschaft.

Fran (France): This research was supported by Assistance Publique des Hôpitaux de Paris (APHP Grant PHRC GAN12), by Institut National de la Santé et de la Recherche Médicale (INSERM grant C0829), by the Fondation FondaMental and by the Investissements d’Avenir Programs managed by the Agence nationale pour la Recherche (references ANR-11-IDEX-0004-02 and ANR-10-COHO-10-01).

Genomic Psychiatry Cohort: The GPC was supported by grants R01 MH085548, R01 MH104964, R01 R01MH104964, and R01MH123451 from the National Institute of Mental Health (NIMH), and genotyping of samples was provided by the Stanley Center for Psychiatric Research at Broad Institute. Funding support for the Whole Genome Association Study of Bipolar Disorder and the Genome-Wide Association of Schizophrenia Study was provided by the NIMH (R01 MH67257, R01 MH59588, R01 MH59571, R01 MH59565, R01 MH59587, R01 MH60870, R01 MH59566, R01 MH59586, R01 MH61675, R01 MH60879, R01 MH81800, U01 MH46276, U01 MH46289, U01 MH46318, U01 MH79469, and U01 MH79470) and the genotyping of samples was provided through the Genetic Association Information Network (GAIN). The following investigators contributed to GPC cohorts:

Michele T Pato MD ^44^, Carlos N Pato, MD, PhD ^44^, Tim B Bigdeli, PhD ^1,2,3^, Ayman H Fanous, MD ^45,46,47^, Steven A McCarroll, PhD ^4,5^, Peter F Buckley, MD ^6^, Mark J. Daly ^7,8,9,5^, James A Knowles MD, PhD ^2,10^, Douglas S Lehrer, MD ^11^, Dolores Malaspina, MD, MSPH ^12,13^, Mark H Rapaport, MD ^14^, Jeffrey J Rakofsky, MD ^14^, Janet L Sobell, PhD ^15^, Giulio Genovese, PhD ^4,5^, Penelope Georgakopoulos, DrPH ^2^, Jacquelyn L Meyers, PhD ^1^, Roseann E Peterson, PhD ^6^, Helena Medeiros, MSW ^2^, Jorge Valderrama, PhD ^1,2^, Eric D Achtyes, MD ^16^, Roman Kotov, PhD ^17^, Colony Abbott, MPH ^16^, Maria Helena Azevedo, PhD ^18^, Richard A Belliveau, Jr, BA ^4^, Elizabeth Bevilacqua, BS ^19^, Evelyn J Bromet, PhD ^17^, William Byerley, MD ^20^, Celia Barreto Carvalho, PhD ^21^, Sinéad B Chapman, MS ^4^, Lynn E DeLisi, MD ^22,23^, Ashley L Dumont, BASc ^4^, Colm O'Dushlaine, PhD ^4^, Laura J Fochtmann, MD ^17^, Diane Gage ^4^, James L Kennedy, MD ^24^, Becky Kinkead, PhD ^14^, Antonio Macedo, PhD ^18^, Jennifer L Moran, PhD ^4^, Christopher P Morley, PhD ^25-27^, Mantosh J Dewan, MD ^27^, James Nemesh ^4^, Diana O Perkins, MD, MPH ^28^, Shaun M Purcell, PhD ^4,29^, Edward M Scolnick, MD ^4^, Brooke M Sklar, MA ^15^, Pamela Sklar, MD, PhD ^12,13^, Jordan W Smoller, MD, ScD ^4,23,30,31^, Patrick F Sullivan, MD, FRANZCP ^28,32^, Humberto Nicolini, MD ^33^, Conrad O Iyegbe, PhD ^34^, Fabio Macciardi, MD, PhD ^35^, Stephen R Marder, MD ^36,37^, Michael A Escamilla, MD ^38^, Ruben C Gur, PhD ^39-41^, Raquel E Gur, MD, PhD ^39-41^, Tiffany A Greenwood, PhD ^42^, David L Braff, MD ^42,43^, Marquis P Vawter, PhD, MA, MS ^35^, Chris Chatzinakos, PhD ^1,2^

^1^ Department of Psychiatry and Behavioral Sciences and ^2^ Institute for Genomics in Health, SUNY Downstate Medical Center, Brooklyn, NY, USA; ^3^ VA New York Harbor Healthcare System, Brooklyn, NY, USA; ^4^ Stanley Center for Psychiatric Research, Broad Institute of MIT and Harvard, Cambridge, MA, USA; ^5^ Department of Genetics, Harvard Medical School, Boston, MA, USA; ^6^ School of Medicine, Virginia Commonwealth University, Richmond, VA, USA; ^7^ Institute for Molecular Medicine Finland (FIMM), University of Helsinki, Helsinki, Finland; ^8^ Analytic and Translational Genetics Unit, Massachusetts General Hospital, Boston, MA, USA; ^9^ Program in Medical and Population Genetics, Broad Institute of Harvard and MIT, Cambridge, MA, USA; ^10^ Department of Cell Biology, SUNY Downstate Medical Center, Brooklyn, NY, USA; ^11^ Department of Psychiatry, Wright State University, Dayton, OH, USA; ^12^ Departments of Psychiatry and ^13^ Genetics & Genomics, Icahn School of Medicine at Mount Sinai, NY, USA; ^14^ Department of Psychiatry and Behavioral Sciences, Emory University, Atlanta, GA, USA; ^15^ Department of Psychiatry & Behavioral Sciences, University of Southern California, Los Angeles, CA, USA; ^16^ Cherry Health and Michigan State University College of Human Medicine, Grand Rapids, MI, USA; ^17^ Department of Psychiatry, Stony Brook University, Stony Brook, NY, USA; ^18^ Institute of Medical Psychology, Faculty of Medicine, University of Coimbra, Coimbra, PT; ^19^ Beacon Health Options, Boston, MA, USA; ^20^ Department of Psychiatry, University of California, San Francisco, CA, USA; ^21^ Faculty of Social and Human Sciences, University of Azores, PT; ^22^ VA Boston Healthcare System, Brockton, MA, USA; ^23^ Department of Psychiatry, Harvard Medical School, Boston, MA, USA; ^24^ Neurogenetics Laboratory, Campbell Family Mental Health Research Institute, Centre for Addiction and Mental Health; Department of Psychiatry, University of Toronto, ON, CA; ^25^ Departments of Public Health and Preventive Medicine, ^26^ Family Medicine, and ^27^ Psychiatry and Behavioral Sciences, State University of New York, Upstate Medical University, Syracuse, NY, USA; ^28^ Department of Psychiatry, University of North Carolina, Chapel Hill, NC, USA; ^29^ Department of Psychiatry, Brigham and Women’s Hospital, Boston, MA, USA; ^30^ Department of Psychiatry, Massachusetts General Hospital, Boston, MA, USA; ^31^ Department of Epidemiology, Harvard T.H. Chan School of Public Health, Boston, MA, USA; ^32^ Medical Epidemiology and Biostatistics, Karolinska Institutet, Solna, SE; ^33^ Carracci Medical Group, Mexico City, MX; ^34^ Department of Psychosis Studies, King’s College London, London, UK; ^35^ Department of Psychiatry and Human Behavior, University of California, Irvine, CA, USA; ^36^ Department of Psychiatry and Biobehavioral Sciences and ^37^ Semel Institute for Neuroscience and Human Behavior, Geffen School of Medicine, University of California Los Angeles, Los Angeles, CA, USA; ^38^ Department of Psychiatry, University of Texas Rio Grande Valley School of Medicine; ^39^ Departments of Psychiatry and ^40^ Child & Adolescent Psychiatry and ^41^ Lifespan Brain Institute, University of Pennsylvania Perelman School of Medicine and Children's Hospital of Philadelphia, Philadelphia, PA, USA; ^42^ Department of Psychiatry, University of California, La Jolla, San Diego, CA, USA; ^43^ VISN-22 Mental Illness, Research, Education and Clinical Center (MIRECC), VA San Diego Healthcare System, San Diego, CA, USA; ^44^ Robert Wood Johnson Medical School, Psychiatry, Rutgers, NJ, USA; ^45^ The University of Arizona College of Medicine-Phoenix; ^46^ Banner-University Medical Center; ^47^ Carl T. Hayden Veterans Administration Medical Center (Phoenix).

Halifax: Halifax data were obtained with support from the Canadian Institutes of Health Research (grant #166098), Genome Canada and from Dalhousie Medical Research Foundation.

The Mayo Bipolar Disorder Biobank was funded by the Marriot Foundation and the Mayo Clinic Center for Individualized Medicine.

Michigan (NIMH/Pritzker Neuropsychiatric Disorders Research Consortium): We thank the participants who donated their time and DNA to make this study possible. We thank members of the NIMH Human Genetics Initiative and the University of Michigan Prechter Bipolar DNA Repository for generously providing phenotype data and DNA samples. Many of the authors are members of the Pritzker Neuropsychiatric Disorders Research Consortium which is supported by the Pritzker Neuropsychiatric Disorders Research Fund L.L.C. A shared intellectual property agreement exists between this philanthropic fund and the University of Michigan, Stanford University, the Weill Medical College of Cornell University, HudsonAlpha Institute of Biotechnology, the Universities of California at Davis, and at Irvine, to encourage the development of appropriate findings for research and clinical applications.

Neuc1 (NeuRA-BDHR-Australia): This work was supported by the Australian National Health and Medical Research Council, grant numbers: 1037196 (PBM, PRS), 1066177 (JMF, JIN), 1063960 (JMF, PRS); and the Lansdowne Foundation. JMF would like to thank Janette M O'Neil and Betty C Lynch for their support.

Neuc1 (NeuRA-CASSI-Australia): This work was funded by the NSW Ministry of Health, Office of Health and Medical Research. CSW was a recipient of National Health and Medical Research Council (Australia) Fellowships (#1117079, #1021970).

Neuc1 (NeuRA-IGP-Australia): MJG was supported by a NHMRC Career Development Fellowship (1061875).

Neuc1/ASGC1/ASRB: This study used samples and data from the Australian Schizophrenia Research Bank (ASRB), which is supported by the National Health and Medical Research Council of Australia, the Pratt Foundation, Ramsay Health Care, the Viertel Charitable Foundation and the Schizophrenia Research Institute. We thank and acknowledge the contribution of the ASRB Chief Investigators: V. Carr, U. Schall, R. Scott, A. Jablensky, B. Mowry, P. Michie, S. Catts, F. Henskens and C. Pantelis.

Span2: CSM was a recipient of a Sara Borrell contract (CD15/00199) and a mobility grant (MV16/00039) from the Instituto de Salud Carlos III, Ministerio de Economía, Industria y Competitividad, Spain. MR was a recipient of a Miguel de Servet contract (CP09/00119 and CPII15/00023) from the Instituto de Salud Carlos III, Ministerio de Economía, Industria y Competitividad, Spain. This investigation was supported by Instituto de Salud Carlos III (PI14/01700, PI15/01789, PI16/01505,PI17/00289, PI18/01788, PI19/00721, and P19/01224), and cofinanced by the European Regional Development Fund (ERDF), “la Marató de TV3” (092330/31), the Agència de Gestió d’Ajuts Universitaris i de Recerca-AGAUR, Generalitat de Catalunya (2014SGR1357 and 2017SGR1461) and the Pla estratègic de recerca i innovació en salut (PERIS), Generalitat de Catalunya (MENTAL-Cat; SLT006/17/287). This project has also received funding from the European Union’s Horizon 2020 Research and Innovation Programme under the grant agreements No 667302 (CoCA) and 728018 (Eat2beNICE).

SWEBIC: We are deeply grateful for the participation of all subjects contributing to this research, and to the collection team that worked to recruit them. We also wish to thank the Swedish National Quality Register for Bipolar Disorders: BipoläR. Funding support was provided by the Stanley Center for Psychiatric Research, Broad Institute from a grant from Stanley Medical Research Institute, the Swedish Research Council, and the NIMH.

Sweden: This work was funded by the Swedish Research Council (M. Schalling, C. Lavebratt), the Stockholm County Council (M. Schalling, C. Lavebratt, L. Backlund, L. Frisén, U. Ösby) and the Söderström Foundation (L. Backlund) and the Swedish Brain Foundation (T. Olsson).

UK - BDRN: BDRN would like to acknowledge funding from the Wellcome Trust and Stanley Medical Research Institute, and especially the research participants who continue to give their time to participate in our research.

UNIBO / University of Barcelona, Hospital Clinic, IDIBAPS, CIBERSAM: EV thanks the support of the Spanish Ministry of Economy and Competitiveness (PI15/00283) integrated into the Plan Nacional de I+D+I y cofinanciado por el ISCIII-Subdirección General de Evaluación y el Fondo Europeo de Desarrollo Regional (FEDER); CIBERSAM; and the Comissionat per a Universitats i Recerca del DIUE de la Generalitat de Catalunya to the Bipolar Disorders Group (2014 SGR 398).

WTCCC: The principal funder of this project was the Wellcome Trust. For the 1958 Birth Cohort, venous blood collection was funded by the UK Medical Research Council.

### **Funding sources**

| **Study** | **Lead investigator** | **Country, Funder, Award number** |
| --- | --- | --- |
| Statistical and functional fine-mapping of bipolar disorder genetic risk loci | N Mullins | USA, Baszucki Brain Research Fund |
| PGC | PF Sullivan; EA Stahl | USA, NIMH MH109528; NIMH U01 MH109536 |
| PGC | D Posthuma | Netherlands, Scientific Organization Netherlands, 480-05-003 |
| PGC | D Posthuma | Dutch Brain Foundation and the VU University Amsterdam Netherlands |
| UK - BDRN | N Craddock  I Jones  L Jones  MJ Owen | Medical Research Council (MRC) Centre (MR/P005748/1G0801418) and Program Grants (MR/P005748/1G0800509) |
| ASRB | V Carr | Australia, National Health and Medical Research Council, grant number: (86500). |
| ASRB | M Cairns | Australia, National Health and Medical Research Council, grant numbers: (1121474, 1147644). |
| ASRB | C Pantelis | Australia, National Health and Medical Research Council of Australia (grants IDs: 1196508, 1150083). |
| BACCS | G Breen | GB, JRIC, HG, CL were supported in part by the NIHR Maudsley Biomedical Research Centre (‘BRC’) hosted at King’s College London and South London and Maudsley NHS Foundation Trust, and funded by the National Institute for Health Research under its Biomedical Research Centres funding initiative. |
| BD_TRS | U Dannlowski | Germany, DFG, Grant FOR2107 DA1151/5-1; Grant SFB-TRR58, Project C09 |
| BiGS, Uchicago | ES Gershon | R01 MH103368 |
| BiGS, NIMH | FJ McMahon | US, NIMH, R01 MH061613, ZIA MH002843 |
| BiGS, GAIN, UCSD | J Kelsoe | US, NIMH, MH078151, MH081804, MH59567 |
| BOMA-Australia | JM Fullerton | Australia, National Health and Medical Research Council, grant numbers: 1066177; 1063960 |
| BOMA-Australia | SE Medland | Australia, National Health and Medical Research Council, grant numbers: 1103623 |
| BOMA-Australia | PB Mitchell | Australia, National Health and Medical Research Council, grant numbers: 1037196 |
| BOMA-Australia | GW Montgomery | Australia, National Health and Medical Research Council, grant numbers: 1078399 |
| BOMA-Australia | PR Schofield | Australia, National Health and Medical Research Council, grant numbers: 1037196; 1063960; 1176716 |
| ROMANIA  rom4_eur  (BOMA-Romania;  Bip_rom3_eur,  bmrom) | M Grigoroiu-Serbanescu | ROMANIA, UEFISCDI, Romania, Grant nr. 203/2021 (code PN-III-P4-ID-PCE-2020-2269) |
| BOMA-Germany I, II, III | S Cichon | Germany, BMBF Integrament, 01ZX1314A/01ZX1614A |
| BOMA-Germany I, II, III | S Cichon | Germany, BMBF NGFNplus MooDS, 01GS08144 |
| BOMA-Germany I, II, III | S Cichon | Switzerland, SNSF, 156791 |
| BOMA-Germany I, II, III | S Cichon | Switzerland, SNSF, 182731 |
| BOMA-Germany I, II, III | MM Nöthen | Germany, BMBF Integrament, 01ZX1314A/01ZX1614A |
| BOMA-Germany I, II, III | MM Nöthen | Germany, BMBF NGFNplus MooDS, 01GS08144 |
| BOMA-Germany I, II, III | MM Nöthen | Germany, Deutsche Forschungsgemeinschaft, Excellence Cluster ImmunoSensation |
| BOMA-Germany I, II, III | MM Nöthen | Germany, Deutsche Forschungsgemeinschaft, NO246/10-1 |
| BOMA-Germany I, II, III | SH Witt | Germany, Deutsche Forschungsgemeinschaft, WI 3429/3-2 |
| BOMA-Germany I, II, III, BOMA-Spain | M Rietschel | Germany, Deutsche Forschungsgemeinschaft, RI 908/11-1 |
| BOMA-Germany I, II, III, BOMA-Spain | M Rietschel | Germany, BMBF, ERA-Net Neuron “EMBED”, 01EW1904 |
| BOMA-Germany I, II, III, BOMA-Spain | M Rietschel | Germany, BMBF, ERA-Net Neuron “Synschiz”, 01EW1810 |
| BOMA-Germany I, II, III, PsyCourse, BiGS | TG Schulze | Germany, BMBF Integrament, 01ZX1314K |
| BOMA-Germany I, II, III, PsyCourse, BiGS | TG Schulze | Germany, DFG, SCHU 1603/4-1, SCHU 1603/5-1, SCHU 1603/7-1 |
| BOMA-Germany I, II, III, PsyCourse, BiGS | TG Schulze | Germany, Dr. Lisa-Oehler Foundation (Kassel, Germany) |
| Bulgarian Trios (Cardiff) | G Kirov  MJ Owen | The recruitment was funded by the Janssen Research Foundation. Genotyping was funded by multiple grants to the Stanley Center for Psychiatric Research at the Broad Institute from the Stanley Medical Research Institute, The Merck Genome Research Foundation, and the Herman Foundation. |
| FOR2107 | T Kircher, U Dannlowski, I Nenadić | German Research Foundation (Deutsche Forschungsgemeinschaft, DFG grant nos. KI 588/14-1, KI 588/14-2, KR 3822/7-1, KR 3822/7-2, NE 2254/1-2, NE 2254/2-1, NE2254/3-1, NE2254/4-1, DA 1151/5-1, DA 1151/5-2, SCHW 559/14-1, 545/7-2, RI 908/11-2, WI 3439/3-2, NO 246/10-2, DE 1614/3-2, HA 7070/2-2, JA 1890/7-1, JA 1890/7-2, MU 1315/8-2, RE 737/20-2, KI 588/17-1) |
| France | M Leboyer, F Bellivier, B Etain, S Jamain | France, APHP, INSERM, ANR, Fondation Fondamental |
| Halifax | M Alda | CIHR grant #166098, Research Nova Scotia, Genome Canada, and Dalhousie Medical Research Foundation |
| IMAGE | B Franke | National Institutes of Health (grant R01MH62873), Dutch NWO Large Investment Program (grant 1750102007010) |
| Mayo Bipolar Disorder Biobank | JM Biernacka, MA Frye | Marriot Foundation and the Mayo Clinic Center for Individualized Medicine |
| Michigan | M Boehnke, RM Myers | US, NIMH, R01 MH09414501A1 |
| Michigan | M Boehnke | US, NIMH, MH105653 |
| Michigan | L Scott | US, NIMH U01 MH085513-01 (sub-contract PI) |
| Mount Sinai, STEP-BD, FAST | P Sklar, EA Stahl | US NIH R01MH106531, R01MH109536 |
| NeuRA-CASSI-Australia (neuc1) | C Shannon Weickert | Australia, National Health and Medical Research Council, grant number: 568807 |
| NeuRA-CASSI-Australia (neuc1) | TW Weickert | Australia, National Health and Medical Research Council, grant number: 568807 |
| NeuRA-IGP-Australia (neuc1) | MJ Green | Australia, National Health and Medical Research Council, grant numbers: 630471, 1081603 |
| NeuRA-BDHR-Australia (neuc1) | PB Mitchell, PR Schofield, JM Fullerton | Australia, National Health and Medical Research Council, grant numbers: 1037196; 1066177; 1063960, 1200428, 1176716, 1177991 & The Lansdowne Foundation |
| Norway | OA Andreassen | Norway, Research Council of Norway (#223273, #248778, #249711, #273291, #296030,#324499 #324252), KG Jebsen Stiftelsen, The South-East Norway Regional Health Authority (#2017-004, #2022-073, #2023-031)  EU’s H2020 RIA grant # 964874 REALMENT |
| Norway | KS O’Connell | US NIH 5R01MH124839-02, Research Council of Norway (#334920) |
| Span2 | M Ribasés | Instituto de Salud Carlos III (PI19/01224, PI20/00041, PI22/00464), “la Marató de TV3” (202228-30 and 202228-31), the Agència de Gestió d’Ajuts Universitaris i de Recerca-AGAUR, Generalitat de Catalunya (2021SGR-00840), Fundació ‘la Caixa, Diputació de Barcelona, Pla Estratègic de Recerca i Innovació en Salut (PERISSLT006/17/285) and Fundació Privada d'Investigació Sant Pau(FISP) |
| State University of New York, Downstate Medical Center (SUNY DMC) | C Pato, MT Pato, JA Knowles, H Medeiros | US, National Institutes of Health, R01MH085542 |
| SWEBIC | M Landén | The Stanley Center for Psychiatric Research, Broad Institute from a grant from Stanley Medical Research Institute, the Swedish Research Council (2022-01643), the Swedish foundation for Strategic Research (KF10-0039), and the Swedish Brain foundation (FO2022-0217). |
| UCL | A McQuillin | Medical Research Council (MRC) - G1000708 |
| UCLA-Utrecht (Los Angeles) | RA Ophoff | US, National Institutes of Health, R01MH090553, R01MH115676 |
| UK - BDRN (Cardiff) | MC O'Donovan | Medical Research Council (MRC) Centre (MR/L010305/1G0801418) and Program Grants (MR/P005748/1G0800509) |
| UK - BDRN (Cardiff) | MJ Owen | Medical Research Council (MRC) Centre (MR/L010305/1G0801418) and Program Grants (MR/P005748/1G0800509) |
| UK - BDRN (Cardiff and Worcester) | N Craddock, I Jones, LA Jones | UK, Wellcome Trust, 078901; USA, Stanley Medical Research Institute, 5710002223-01 |
| UK - BDRN (Cardiff) | A Di Florio | European Commission Marie Curie Fellowship, grant number 623932. |
| UNIBO / University of Barcelona, Hospital Clinic, IDIBAPS, CIBERSAM | E Vieta | Grants PI15/00283 (Spain) and 2021 SGR 01358 (Catalonia) |
| University of Pittsburgh | V Nimgaonkar | US, NIMH MH63480 |
| USC | JL Sobell | USA, National Institutes of Health, R01MH085542 |
| WTCCC | N Craddock; AH Young | Wellcome Trust. For the 1958 Birth Cohort, venous blood collection was funded by the UK Medical Research Council. AHY was funded by NIMH (USA); CIHR (Canada); NARSAD (USA); Stanley Medical Research Institute (USA); MRC (UK); Wellcome Trust (UK); Royal College of Physicians (Edin); BMA (UK); UBC-VGH Foundation (Canada); WEDC (Canada); CCS Depression Research Fund (Canada); MSFHR (Canada); NIHR (UK); Janssen (UK) |
| Greece | G.P. Patrinos | European Commission (H2020-668353; Ubiquitous Pharmacogenomics (U-PGx); European Commission (Horizon Europe-101057639; SafePolyMed); Greek General Secretariat of Research and Technology (MIS 5002550) |
| University of California, Irvine | MP Vawter | USA, National Institutes of Health, MH113177, MH074307, MH085801, MH099440, RR000827, MH060068, MH060870.  Pritzker Neuropsychiatric Disorders Research Fund L.L.C. |
| Korea | HJ Lee,  Woojae Myung, Hong-Hee Won | National Research Foundation of Korea (2019R1A2C2084158 and 2017M3A9F1031220 to HJ Lee; NRF-2021R1A2C4001779 to WM;NRF-2022R1A2C2009998 to HHW). Ministry of Health & Welfare of Korea (HM14C2606) |
| BIPGRAZ, Austria | EZ Reininghaus | Funding from the government of Styria, Austria. |
| BIPGRAZ, Austria | SA Bengesser | MEFO funding of the Medical University of Graz. |
| Japan | N Iwata, M Ikeda | Jakpan Agency for Medical Research and Development  (JP20dm0107097, JP20km0405201, JP20km0405208) |
| FAST-STEP | JW Smoller | USA, NIH, R01MH063445 |
| ICCBD | JW Smoller, P Sklar | USA, NIH, R01MH085542 |
| Edinburgh, Scotland | A McIntosh | Wellcome funding, reference 220857/Z/20/Z |
